# Influence of intestinal schistosome and hepatitis B or C coinfection on hepatic disease: a systematic review and meta-analysis

**DOI:** 10.1101/2025.04.10.25325597

**Authors:** Laura Kmentt, Lauren Wilburn, Huike Cheng, Goylette F. Chami

## Abstract

**Background:** Intestinal schistosome and hepatitis B and C infections independently cause liver disease. Yet, there is no consensus on the relative influence of schistosome and viral hepatitis coinfection for any liver disease.

**Methods:** We conducted a meta-analysis for intestinal schistosome and hepatitis B or C coinfection on author-defined nonspecific liver outcomes, liver fibrosis, cirrhosis and hepatocellular carcinoma. The study protocol was prospectively registered on PROSPERO (CRD42023443435) and adhered to PRISMA reporting guidelines. The Cochrane Central Register of Controlled Trials, Embase, Global Health, Global Index Medicus, and Medline were systematically searched from inception to 23 January 2025. Inverse-variance weighted random effects were used to calculate pooled effect sizes. Subgroup analyses were conducted for study aims, region, species, diagnostic tools, and reference categories of singularly infected versus uninfected. We assessed study quality using a modified National Institute of Health risk of bias (RoB) tool.

**Findings:** Out of 1984 studies screened, 33 full text articles were eligible for meta-analysis with 8637 participants. 57% of studies (19/33) were on coinfection with hepatitis C and *S. mansoni.* Individuals with any coinfection were 2·75 times more likely to have any liver disease than singularly or uninfected individuals, and a 2·29, 2·35 and 2·69 higher likelihood of liver fibrosis, cirrhosis, and hepatocellular carcinoma, respectively. Schistosome and hepatitis B coinfections, in particular, were 4·11 times more likely to be associated with any liver disease. Results were similar when compared to singularly infected only. Heterogeneity was moderate (I^2^ 74·35%), and 42·42% (14/32) of studies where RoB could be assessed were of low quality.

**Interpretation:** Schistosome and viral hepatitis coinfection worsened hepatic disease. Guidelines for schistosomiasis and hepatitis B and C should consider coinfection when evaluating eligibility for treatment or prophylaxis, and determining morbidity management strategies.

**Funding:** NDPH Pump Priming Fund, John Fell Fund, Robertson Foundation, and UKRI EPSRC (EP/X021793/1).

**Research in context:** *Evidence before the study:* Schistosomiasis, caused by helminths of the species *Schistosoma mansoni, S. japonicum, S. mekongi,* as well as chronic infection with hepatitis B and C viruses can lead to similar liver disease outcomes including periportal or portal fibrosis. In 2024, the World Health Organization (WHO) released new guidelines for hepatitis B that changed treatment eligibility based on the importance of coinfections, of which schistosomiasis was not considered despite known co-endemicity. For schistosomiasis, there are currently no WHO guidelines that directly focus on morbidity management. A population-based study in Uganda by Anjorin and colleagues showed that schistosomal liver fibrosis risk depended on underlying hepatitis B coinfections. However, there is conflicting evidence as to the association of intestinal schistosome and hepatitis B or C coinfections, especially given the current context of available hepatitis B vaccinations and regular mass drug administration for schistosomiasis. It remains unknown how similar the hepatic disease presentations are between these vastly different helminth and viral pathogens; whether diseases specific to one pathogen can be worsened by coinfection; or whether an individual could be predisposed to develop hepatic disease if exposed previously to the other pathogen. These knowledge gaps exist despite known spatial overlap in the portal area of liver fibrosis caused by both pathogens. There remain open questions as to potential interactions between immune-driven inflammatory processes specific to schistosomes versus hepatitis B or C, or any role of immune priming for liver fibrosis. Here we synthesised the current state of evidence to assess whether coinfection worsens hepatic outcomes when compared to singular or no infections and to identify the relevance and severity of the type of hepatic outcome in humans only with murine models excluded. The Cochrane Central Register of Controlled Trials (1996-), Medline including PubMed (1946-), Embase (1974-), Global Health (1973-) and Global Index Medicus (1901-) were searched from database inception to 7 July 2023 and updated on 23 January 2025 using the search string (schistosom* OR bilharzia* OR "snail fever" OR “mansoni” OR “japonicum” OR “mekongi”) and (“hepatitis B” OR HBV OR “hepatitis C” or HCV or “hepatitis B C”) AND (liver OR hepat* OR cirrho*). One systematic literature review was identified from this search string, which summarised the clinical progression of liver disease in general in the event of coinfection of *S. mansoni* and hepatitis B or C. However, this review did not perform a meta-analysis and was focused solely on coinfection relating to one species of intestinal schistosomiasis. We found no published reviews specifically investigating coinfection with any intestinal schistosome species and hepatitis B or C for hepatomegaly, liver fibrosis, cirrhosis and hepatocellular carcinoma.

*Added value of this study:* In this systematic review and meta-analysis, we estimated the pooled effect size of coinfection versus singular infections or no infection, as well as compared to singular infections only. We assessed whether intestinal schistosomes and viral hepatitis B or C coinfections influence the odds of nonspecific liver pathology, liver fibrosis, cirrhosis and hepatocellular carcinoma without restrictions on language or geography. By combining summary measures from 8637 participants across 7 different countries from 1991-2024, we identified that co-infected individuals were 2·75 times more likely to have any liver pathology than singularly or uninfected individuals, and the odds were similar when coinfected individuals were compared to singularly infected individuals only (Odds Ratio 2·61; CI 1·62-4·25). When specified by author-defined hepatic disease outcomes, we found that coinfection was over two times more likely to be associated with liver fibrosis, cirrhosis, and hepatocellular carcinoma when compared to singularly or uninfected individuals. The results were similar when compared to singularly infected individuals only, though slightly attenuated for hepatocellular carcinoma (Odds Ratio 1·81; CI 1·11-2·96). Remarkably, schistosome and hepatitis B coinfection had over four times higher likelihood of any hepatic outcome than singular infections. Heterogeneity amongst the included studies was moderate (I^2^ 74·35%) and was reduced (I^2^ 69·85%) when outlying studies were removed. Risk of bias was moderate to high in most included studies, with only one study classed as low risk of bias.

*Implications of all the available evidence:* This study demonstrates the importance of jointly considering schistosomes and hepatitis B or C for estimating the likelihood of chronic liver diseases of varying prognostic value. Coinfections influenced a range of author-defined hepatic outcomes of varying severity from fibrosis to cirrhosis. Future research is needed to assess whether to incorporate coinfections in guidelines for schistosomiasis and hepatitis B or C morbidity management, as well as to explore possibilities of coordinating vaccination campaigns with mass drug administration, or preventative interventions such as health education targeting.

## Introduction

An estimated 1 in 25 deaths worldwide—over 2 million annually—are due to a liver-related disease.^1^ Complex aetiologies with both infectious and non-communicable causes can lead to common liver pathologies including liver fibrosis, cirrhosis and hepatocellular carcinoma (HCC).^2–6^ Two leading causes of liver disease in low-income countries, in particular as related to liver fibrosis, are intestinal schistosomiasis (caused by helminths of the species *Schistosoma mansoni, S. japonicum,* and *S. mekongi*) and viral infections with hepatitis B (HBV) and C (HCV). Schistosome infection occurs through human contact with parasite cercariae in freshwater with competent intermediate snail hosts, whereas HBV and HCV are transmitted through bodily fluid contact or vertical transmission from mother to child.^2,7,8^

Despite their varied routes of transmission, the geographical areas as well as populations at risk of these infections overlap.^9^ Global Burden of Disease (GBD) data estimates 12,900 deaths were caused by schistosomiasis globally in 2021, of which 83·9% occurred in sub-Saharan Africa.^10^ Countries with the highest mortality rates due to schistosomiasis included the Central African Republic, Democratic Republic of Congo, Somalia, Zimbabwe, Congo, Mozambique and Burundi.^10^ Global HBV infection prevalence is estimated at around 3·2%, with great variability between countries and regions, with some Sub-Saharan African countries such as Niger with a HBsAg positivity prevalence of 16·2%, or 18·7% in Guinea Bissau.^11^ Globally, it is estimated that the age-standardised death rate for cirrhosis caused by HBV was 4·03 per 100,000 population, exceeding 10 per 100,000 population in most sub-Saharan African countries, as well as several Asian countries.^12^ Highest all-age mortality rates from chronic HBV in Africa in 2021 were found in Egypt, Central African Republic, Congo, Kenya, South Sudan, Somalia, Eritrea, Chad, Guinea-Bissau and Malawi.^10^ Egypt, Central African Republic, Congo and Kenya are among the most affected countries in Africa in terms of all-age mortality from chronic HCV.^10^ However, mortality rates are grossly underestimated given any comorbidity or shared causes are not considered in GBD estimates.

Coinfection is well established and common among schistosomes and HBV or HCV, although studies have concentrated on high burden countries for HBV or HCV, and patients within healthcare facilities.^13–16^ Amongst adult patients admitted to healthcare facilities in Brazil, the prevalence of *S. mansoni* and HBV was found to range between 30-44%^9,17,18^ and 12·9% for coinfection with HCV.^19^ In China, HBV infection amongst adults with schistosomiasis was found to range between 25·37% to 58·4% in rural populations and patients attending hospitals for liver-related pathologies, respectively.^13,20^ In Egypt, which remains one of the countries with the highest burden of HCV infection globally, ^10^the prevalence of coinfection with *S. mansoni* has been demonstrated to be 33% amongst adult liver disease patients,^21^ and coinfection with HBV has been estimated at 19·6% amongst adults and children in the community.^15^

The potential shared mechanisms of pathogenesis for schistosomes and HBV or HCV are unclear, despite spatial overlap of the liver fibrosis development in the portal area depending on the stage of schistosomal fibrosis. Liver damage from schistosome infection is a result of immune-modulated fibrogenesis—a granulomatous response—to egg antigens in the portal venules.^22^ Repeated exposure, or an uncontrolled or excessive Th2 response may result in the granulomatous-driven fibrosis progressing to the adjacent (periportal) parenchyma. Only rarely does persistent or repeated fibrosis progress to cirrhosis despite producing end-stage hepatic complications such as portal hypertension.^2,4^ Few studies, focused on *S. japonicum,* suggest that schistosome infection may be associated to repeated hepatocyte damage and, in turn, the development of liver malignancy.^4,23,24^ Yet, murine studies suggest hepatocyte death is rare, at least certainly in the case on in-tact granulomas caused by *S. mansoni.*^25^ Liver parenchymal damage in viral hepatitis infection occurs through virus entry into hepatocytes and replication, resulting in an immune-mediated inflammatory response and hepatocyte damage, and subsequent attempts at hepatocellular repair through fibrogenic and inflammatory processes.^5,6,26^ Chronic viral hepatitis usually initially leads to fibrotic changes in the periportal tract, later progressing to bridging fibrosis and eventually frank cirrhosis.^26^ Persistent infection with repeated cycles of hepatocyte repair, can lead to chromosomal damage resulting in malignant transformation to HCC, as well as progressive expansion of fibrosis and eventual liver cirrhosis.^5,6,27^ Yet, despite differences in pathogenesis, coinfection with intestinal schistosomes and HBV and HCV has been shown to worsen clinical outcomes through enhancing immune-mediated hepatocyte damage, enabling hepatitis virus entry into hepatocytes as well as prolonging the HBV carrier state leading to increased hepatitis-driven liver pathology, although there is no consensus in the existing literature.^18,28,29^

While prevalence of schistosome and HBV or HCV coinfection, albeit with highly varied results, has been extensively studied in endemic settings, it remains an open question as to whether and how coinfection influences liver pathology. Without restrictions for language or geography, we estimated the pooled effect size of schistosome and HBV or HCV coinfection versus singular infections or no infection, as well as compared to singularly infections only, for the odds of nonspecific liver pathology, liver fibrosis, cirrhosis and HCC.

## Methods

The study protocol was registered on 10 July 2023 on PROSPERO (CRD42023443435). The Cochrane Central Register of Controlled Trials (1996-), Medline including PubMed (1946-), Embase (1974-), Global Health (1973-) and Global Index Medicus (1901-) were searched from database inception to 7 July 2023 using the search string (schistosom* OR bilharzia* OR "snail fever" OR “mansoni” OR “japonicum” OR “mekongi”) AND (“hepatitis B” OR HBV OR “hepatitis C” or HCV or “hepatitis B C”) AND (liver OR hepat* OR cirrho*). The search was updated on 23 January 2025 using the same search string. See Appendix 1 pp4-6 for the full search string and database hits. References were exported to Covidence software^30^ for de-duplication with manual removal of any missed duplicates and for later screening by two reviewers.

Studies set in non-endemic areas, not on humans, on hepatitis species other than B and C or on schistosome species other than *S. mansoni*, *S. japonicum* and *S. mekongi* were excluded. There were no exclusions by language, geography, or participant characteristics. Only studies where viral hepatitis was diagnosed by antigen, antibody, or viral load were eligible for inclusion. Similarly, diagnosis of schistosomiasis was limited to antigen, antibody, or microscopy-based tools. Studies using hospital records for schistosomiasis diagnosis were also included. Eligible liver morbidity diagnostics included biopsy (for fibrosis, cirrhosis and HCC), imaging (for hepatomegaly, fibrosis, cirrhosis and HCC) and biomarkers (when specifically characterised as being utilised for fibrosis, cirrhosis or HCC diagnosis). See Appendix 1 p7 for full inclusion and exclusion criteria.

Abstracts and full text reports were independently screened by LK and LW. Eligibility conflicts were resolved between the two reviewers. Included studies were extracted by LK using a pre-validated extraction template, and 10% of extracted studies were independently cross checked by LW as a quality control measure. Where effect sizes were not reported by study authors, odds ratios (ORs) where reconstructed using in-text information when possible. Attempts were made to contact the corresponding authors of the study via email to obtain information to construct ORs if not already provided in text. Studies with insufficient information to extract effect sizes, and where contacting study authors had failed after two attempts, were excluded. Studies where full text articles could not be obtained by independent searches online or by requesting texts from the Bodleian Libraries at the University of Oxford or the British Library, were excluded. Non-English studies were first screened by LW using an online translation software. Through this process, we identified three studies in Mandarin Chinese that were subsequently reviewed by a native speaker, HC, for possible inclusion based on our pre-defined inclusion and exclusion criteria. Two of these studies were included in the meta-analysis and extracted by HC and LK. A full list of excluded studies with reasons for exclusion can be found in Appendix 1 pp8-16.

The exposure of interest, active or past coinfection with one or multiple *Schistosoma* species and HBV or HCV, was extracted based on the author-provided definitions of the level of infection and coded as a binary variable. Depending on the study, the reference category could have been singularly infected, uninfected, or both singularly and uninfected individuals. The outcomes and ORs of any presence of hepatomegaly, liver fibrosis, cirrhosis or HCC, defined by authors, were extracted if available. We also extracted information on study design including country, region, study setting, participant sex and ages, study duration, sampling strategy, inclusion and exclusion criteria, study aim and diagnostic modality used for diagnosis of exposure and outcome (see Appendix 1 pp17-20 for a list of all extracted variables).

All statistical analyses were completed in Stata MP v17. Random effects models were used to calculate pooled effect sizes (ORs and 95% confidence intervals (CIs)) of coinfection for specific liver outcomes as well as a nonspecific outcome of any liver disease. The non-specific outcome of any liver disease was constructed from the other three liver pathology outcomes: if a study provided one liver pathology outcome, such as fibrosis, this was used. If a study provided more than one outcome, such as reporting on fibrosis and cirrhosis, the more severe outcome, in this case cirrhosis, was chosen. The weight of each study was calculated using the inverse sum of within-study variance and between-study variance. The Higgin’s I^2^ statistic was used to inspect heterogeneity between studies. If three or more comparable studies were available for any of the extracted study variables, subgroup analyses were performed. Subgroups included study aim, hepatitis species, *Schistosoma* species, World Bank region, diagnostic modality, risk of bias and the aforementioned subgroups rerun with studies using a reference category of singularly infected only. Subgroups were also rerun excluding studies with a high risk of bias.

Each included study was assessed for risk of bias (RoB) using a modified version of the Quality Assessment Tools for observational cohort and cross-sectional studies, case control studies and before-after studies with no control group by the National Institutes of Health (see Appendix 1 pp21-35 for full risk of bias assessment). Funnel plots and Egger’s test were used to assess publication bias (see Appendix 1 p36).

## Results

The study selection flowchart is presented in Figure 1. The search returned 5578 records. Following removal of duplicates, 1984 unique titles and abstracts were screened with 33 full text articles eligible for meta-analysis.^14,31,40–49,32,50–59,33–39^ The most common reason for exclusion at the full text stage (208 articles excluded) was due to ineligible outcomes being studied (126 articles), such as prevalence of coinfection, or no coinfection as an exposure.

**Figure 1:**
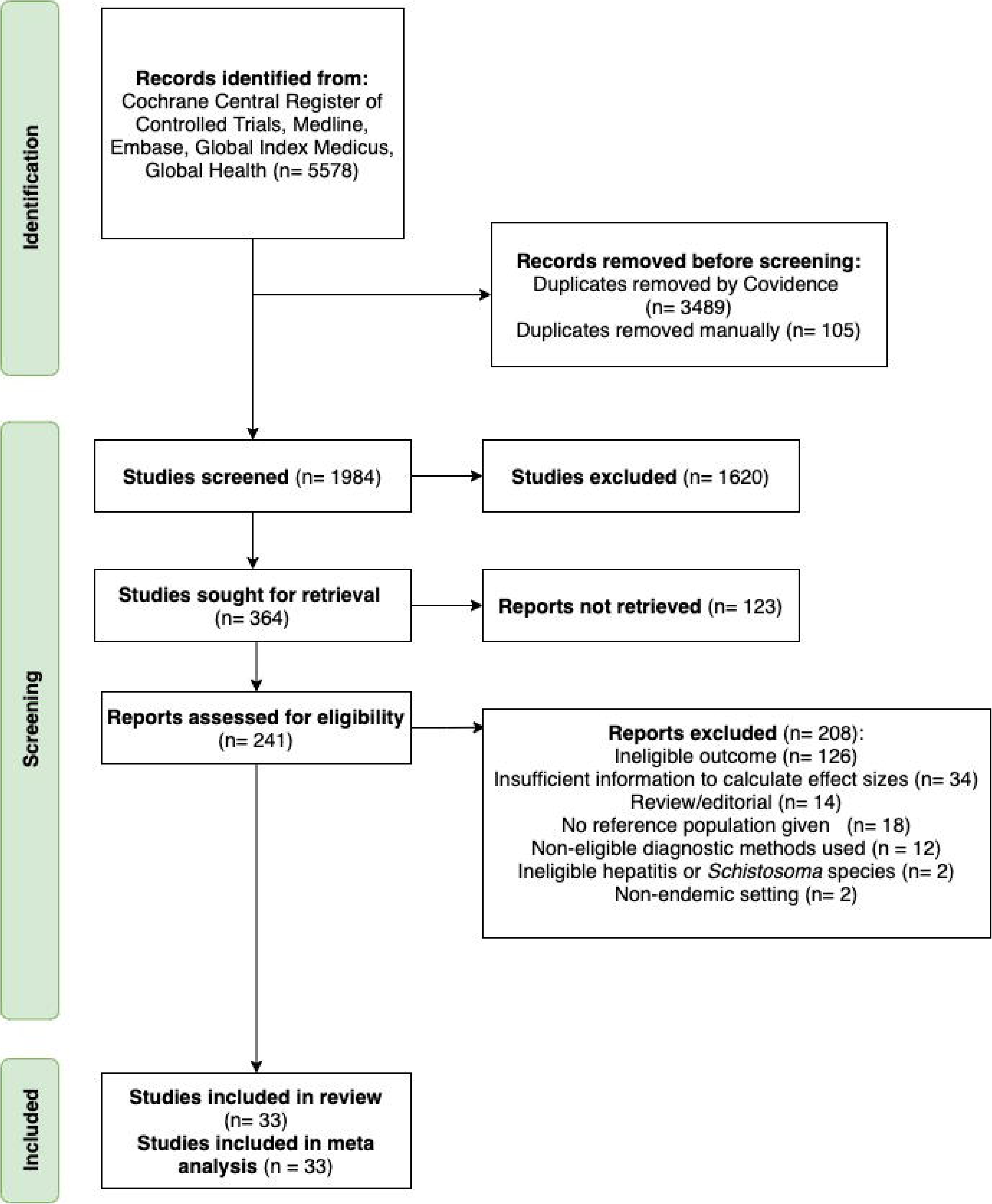
PRISMA flowchart of study selection.

Infection and coinfection characteristics as well as liver outcomes are summarised in Table 1. The total number of participants across all 33 studies was 8637. The most commonly studied schistosome species was *S. mansoni* (24/33, 72·73%)^31,32,42–46,48,50–52,54,33,55,60–62,34–36,38–41^, followed by *S. japonicum* (7/33, 21·21%).^37,47,53,56–59^ Two studies reported on combined *S. mansoni* and *S. haematobium* coinfection (2/33, 6·06%)^14,49^. No included studies reported on *S. mekongi.* HCV (22/33, 66·67%)^31,32,41–46,49,51,54,55,33,56,60,34–40^ was more commonly studied than HBV (5/33, 15·15%)^47,48,50,57,59^ or mixed B and C analyses (6/33, 18·18%).^14,52,53,58^ The most common coinfection observed was *S. mansoni* with HCV (19/33, 57·00%).^31,32,42–46,51,54,55,60,33–36,38–41^ In all but ten studies (23/33, 69·69%),^31,32,42,43,48–51,53–56,33,57–61,35–41^ the reference population was singularly infected individuals. 14/25 (56·00%) had a reference population of singularly infected with HCV^31,33,51,54,60,35,36,39–43,49^ or HBV^50^ , 3/25 (12·00%) had a reference population of singularly infected with *S. japonicum*^57,59^ or *S. mansoni*,^55^ and 7/25 (28·00%)^37,38,45,48,53,56,61^ had a mixed reference population with singular schistosomiasis or singular hepatitis infection. Ten studies (10/33, 30·30%)^14,32,34,44–47,52,58,62^ mixed singularly infected and uninfected participants and healthy controls to form a reference group. No study had a completely uninfected reference group.

**Table 1:**
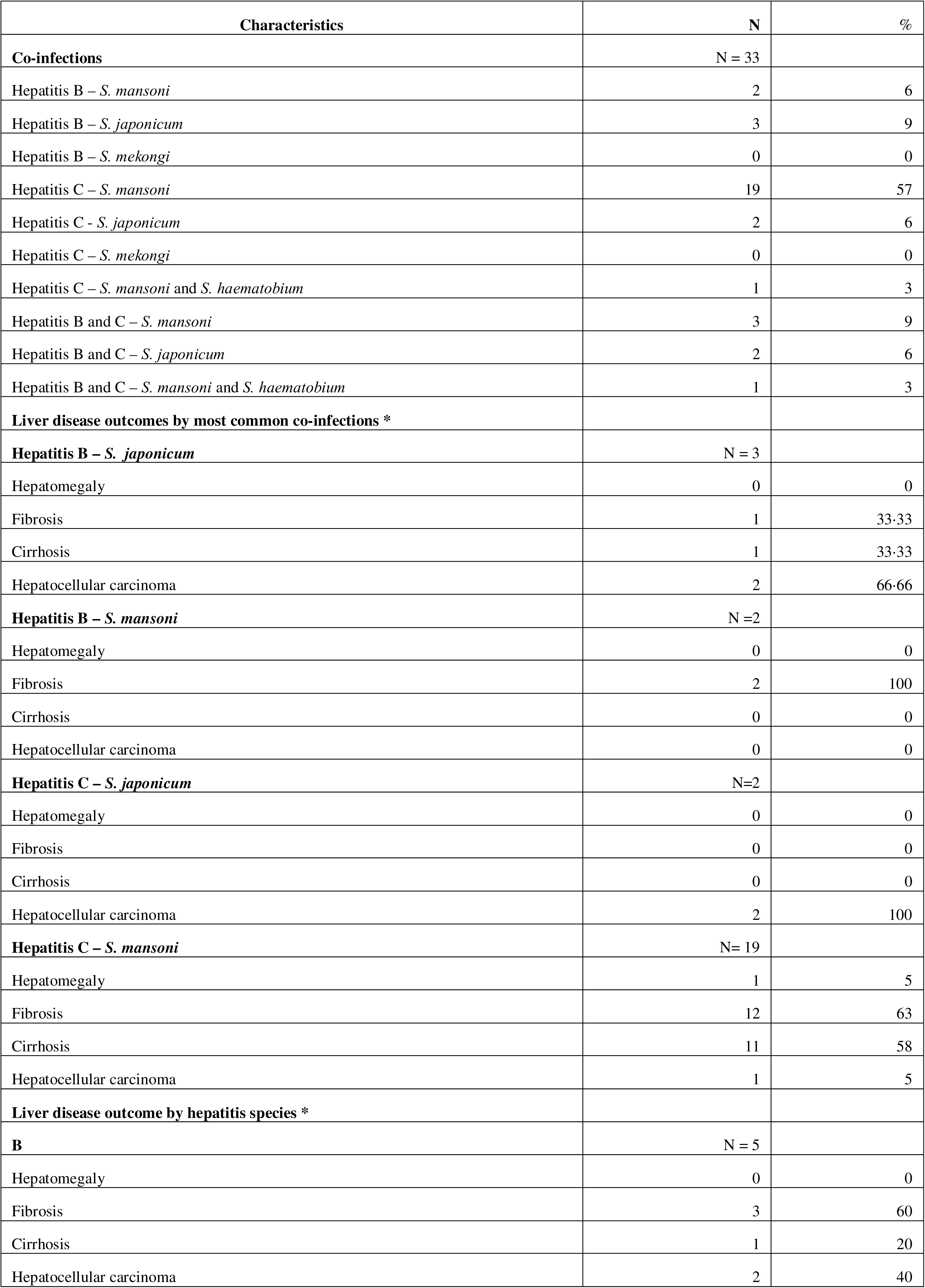
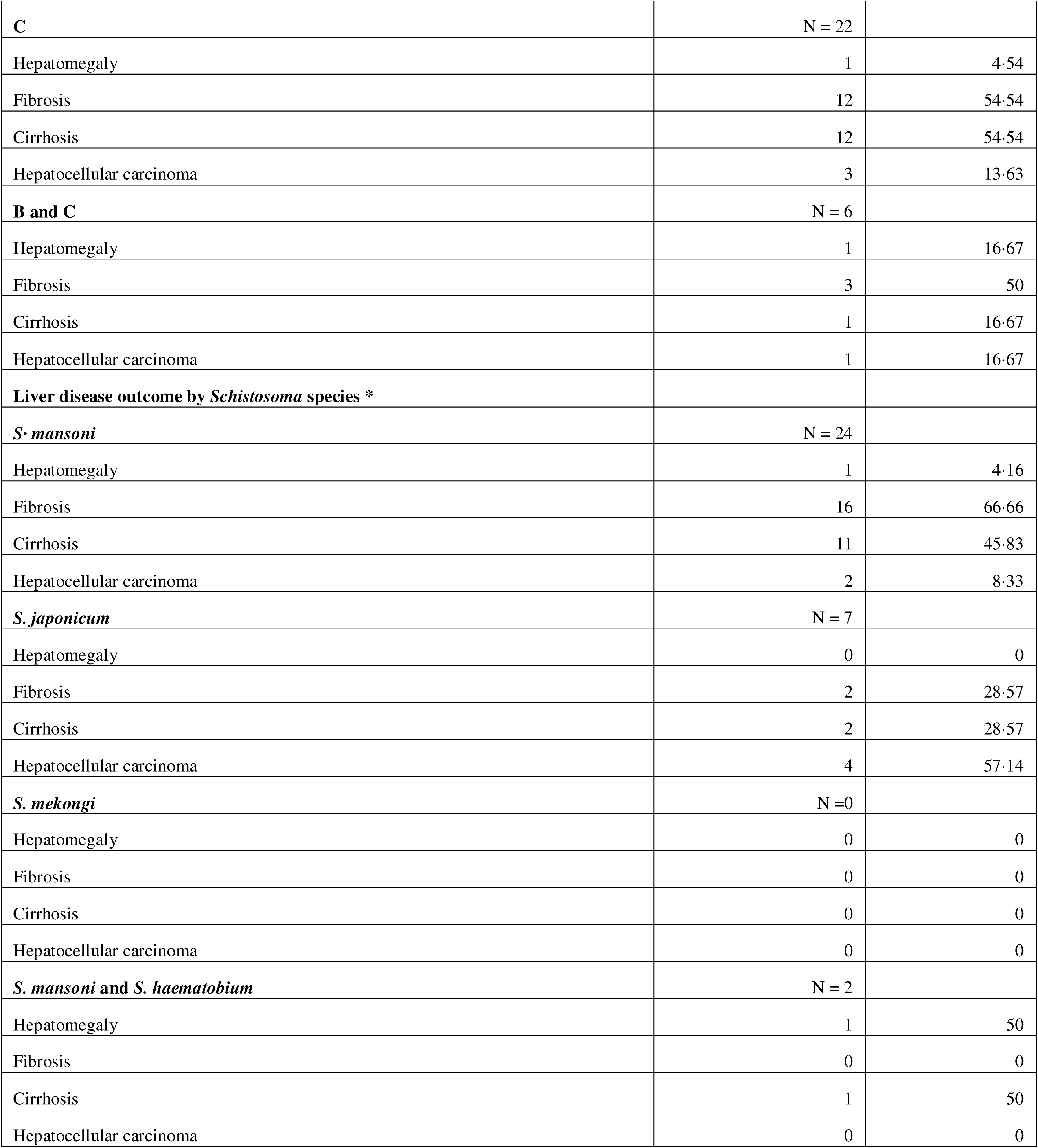
Summary of infection and coinfection characteristics across all included studies. *Liver pathologies listed by coinfection are not mutually exclusive (i.e. a study focusing on coinfection between hepatitis B and *S. mansoni* can report on both fibrosis and cirrhosis as liver disease outcomes.

18 of 33 studies (54·54%)^31,32,48,50–53,55,60,62,33,35,36,39,40,45–47^ focused on liver fibrosis, 14 (42·42%) on cirrhosis,^31,33,54,55,58,59,34–36,38,41–43,49^ 6 (18·18%) on HCC^37,44,56–58,61^ and 2 (6·06%) on hepatomegaly.^14,40^ The most common diagnostic modality for fibrosis was biopsy (8/18, 44·44%)^31,32,35,36,39,46,51,60^ and imaging findings (7/18, 38·89%).^33,40,47,50,52,55,62^ Cirrhosis was most commonly diagnosed by biopsy (8/14, 57·14%)^31,35,36,41–43,49,59^ followed by imaging (4/14, 28·57%). ^33,34,54,55^ HCC was mostly diagnosed by biopsy (3/6, 60.00%).^37,56,57^ The two studies reporting on hepatomegaly^14,40^ did not state the exact method of diagnosis.

Study characteristics are provided in Table S13 in Appendix 1 pp37-39. 32 of 33 studies (96·97%)^31,32,41–50,33,51–60,34,61,62,35–40^ were on both males and females, assuming that studies where the sex of participants was not mentioned (2/33) had included both males and females. Studies most commonly aimed to investigate pathogenesis in coinfection (14/33, 42·42%)^31,34,58,59,61,62,35–37,49,53,55–57^ followed by studies on immune response to coinfection (5/33, 16·67%)^32,33,38,39,51^ and studies investigating specific biomarkers of liver damage (6/33, 18·18%).^43–46,52,60^ Most studies were set in Middle East/North Africa (18/33, 54·55%).^14,31,43,44,46,49,54,55,60,61,32–36,38,41,42^ Four studies (12·12%)^40,48,52,62^ were set in sub-Saharan Africa and seven studies (21·21%)^37,47,53,57–59^ were set in East Asia/Pacific. Most _studies (22/34, 64·70 %)_^31,32,44,46,48,49,53–56,58,59,33,60,61,34,35,37–39,41,43^ were based in hospitals with only 5 out of 34 (14·70%) ^14,40,52,58,62^ in community health centres and 7 out of 33 (20·59%)^36,42,45,47,50,51,57^ had unclear study settings. One study^58^ was conducted in both community and hospital settings. The most common study design was cross-sectional (21/33, 63·64%)^14,33,46,47,52,54–59,61,34,62,35,36,40–42,44,45^, followed by prospective cohort (7/33, 21·21%)^31,32,38,39,43,51,60^ and retrospective case control (3/33, 9·09%).^48–50^

Across all studies, 30·30% (10/33)^32,35,62,38,41–43,46,49,51,61^ included patients based on serological findings; only one study (3·03%)^48^ included participants based on imaging findings whereas, most studies (13/33, 39·39%)^14,31,55–57,33,34,37,45,47,50,52,54^ did not provide any specific explanation on the selection criteria for their study participants. Patients were mostly excluded (13/33, 39·39%)^36,38,55,60,61,39–42,44,46,49,51^ based on the presence of liver comorbidities such as autoimmune hepatitis,^31^ alcoholic liver disease^38^ or alpha 1-antitrypsin deficiency.^41^ The most common diagnostic modality used for *Schistosoma* diagnosis was microscopy (22/33, 66·67%),^14,31,44–48,51–53,55,59,32,61,62,34,35,37–40,42^ followed by serology (18/33, 54·55%) ^32,33,50,51,53–55,58,60,61,35,36,39,41,43,45,46,49^ and other methods such as diagnoses extracted from medical records or biopsy (13/33, 39·39%)^34,35,56,57,36–39,45–47,51^ and imaging (3/33, 9·09%).^34,37,59^ The methods given are not mutually exclusive and multiple diagnostic methods were used across studies.

Individuals with any coinfection between intestinal schistosomes and HBV or HCV had 2·75 times higher odds of any liver disease, i.e. either liver fibrosis, cirrhosis, or HCC, when compared to individuals with singular infections or no infections (Figure 2).

**Figure 2:**
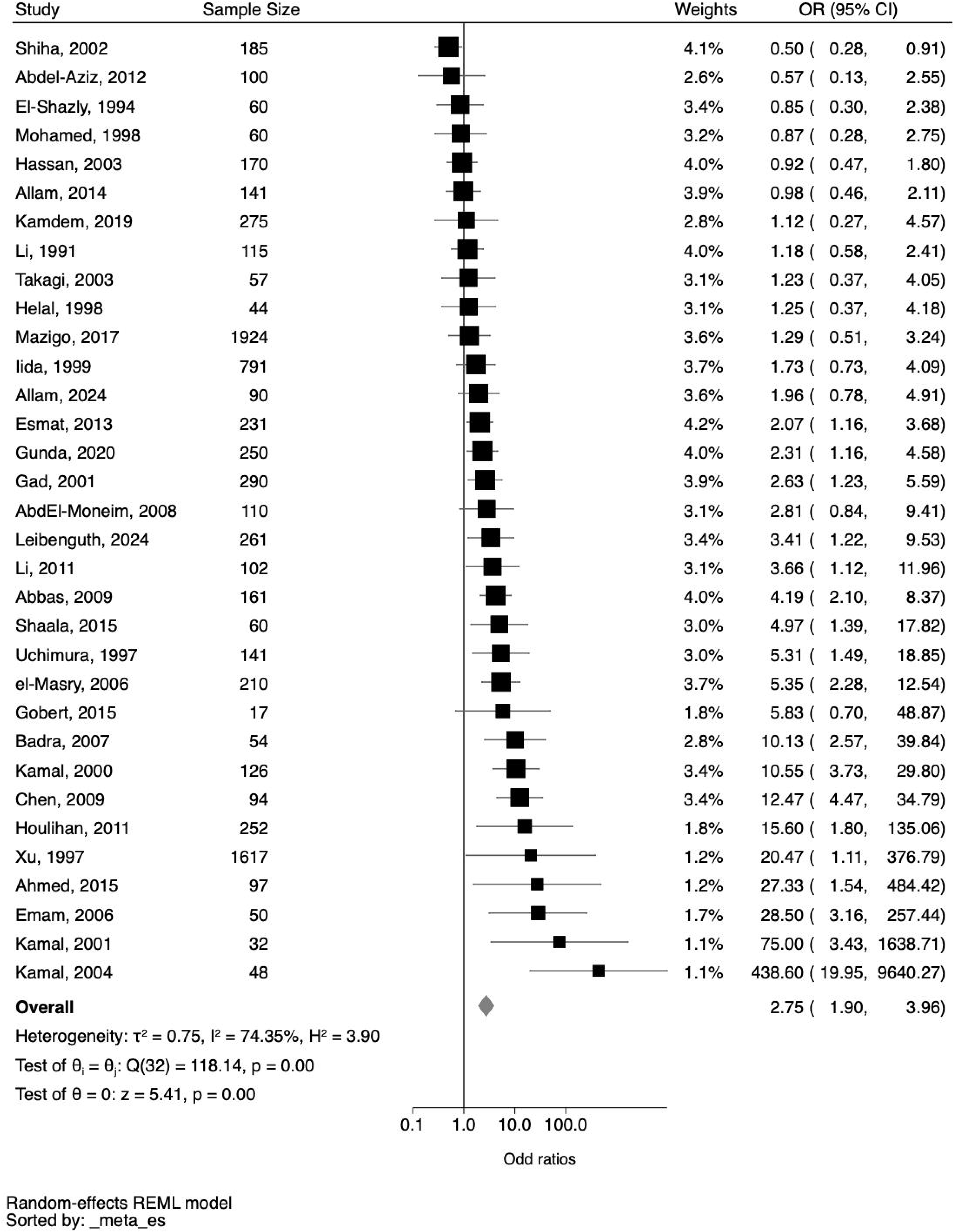
Pooled effect size of coinfections undifferentiated by the type of liver disease based on 33 studies. OR (Odds Ratio), 95% CI (95% Confidence Interval), I^2^ = statistic of heterogeneity.

Heterogeneity for this model was moderate (I^2^ 74·35%) and was reduced when outliers^39,46,51,60^ (studies with ORs greater than 25) were removed (I^2^ 69·85%) (see Figure S2 Appendix 1 p40). For liver fibrosis, there was a 2·29 times higher odds (CI 1·46-3·58) of liver fibrosis in coinfected individuals than in singularly infected or uninfected individuals (Figure S3 Appendix 1 p41) with moderate heterogeneity (I^2^ 64·02%), which was reduced to 61·03% when outliers with very high ORs^39,51,60^ were removed (Figure S4 Appendix 1 p42). Coinfection was associated with a 2·35 times higher odds of liver cirrhosis compared to single infection or no infection (CI 1·37 – 4·02) with moderate heterogeneity (I^2^ 68·31%) (Figure S5 Appendix 1 p43). Similarly, individuals with schistosome and HBV or HCV coinfections were 2·69 times as likely to have HCC than individuals with single infections or no infection (CI 1·45 – 4·99) with moderate heterogeneity (I^2^ 65·38%) (Figure S6 Appendix 1 p44). Only two studies^32,48^ reported adjusted data for liver fibrosis. No cirrhosis or HCC studies reported adjusted data. Analyses were rerun excluding ten ^14,32,34,44–47,52,58,62^, six,^32,45–47,52,62^ two,^34,58^ and one^58^ studies that included uninfected individuals as part of the reference population for nonspecific liver outcomes, liver fibrosis, cirrhosis, and HCC, respectively. One study on hepatomegaly^14^ containing controls was also excluded. All results remained robust and of similar magnitude when coinfection was compared to singular infections and OR attenuation was observed only for HCC (OR 1·81; 1·11 – 2·96, I^2^ 16·59%) (see Table S14 Appendix 1 p45).

Subgroup analyses for any liver pathology are shown in Table 2. Fibrosis and cirrhosis subgroup analyses are provided in Appendix 1 pp46-47 (Text S2, Table S10, S11). Coinfection influences were greatest in studies focusing on immune responses, although with high variability (OR 11·41, CI 1·65 - 78·78) followed by studies on biomarkers (OR 3.46, CI 1·14 – 10·49) then studies of pathogenesis (OR 2·38, 95% CI 1·45- 3·92). No significant effects within biomarker studies were found after removing studies that included uninfected individuals in the reference group (Table 2). For liver fibrosis, the strong association with the aim of studying immune responses was lost and turned insignificant (Table S15 Appendix 1 p47). Coinfection with *S. japonicum* and HBV or HCV showed somewhat stronger associations with any liver disease (OR 3·76, CI 1·74 - 8·14) than with *S. mansoni* and HBV or HCV (OR 2·80, CI 1·78 - 4·40), remaining robust in studies with only singularly infected individuals as the reference population (Table 2). The effect of HBV (OR 4·11, CI 1·46 - 11·59) and any schistosome coinfection on the development of any liver pathology was greater when compared to HCV (OR 2·78, CI 1·70 - 4·54), but insignificantly different with fewer studies on HBV. Where only singularly infected individuals were included as a reference population or the outcome was liver fibrosis, results remained similar (Table 2). Studies using biopsy as their diagnostic modality, found an association between coinfection and the development of fibrosis (OR 3·12, CI 1·02 – 9·50) but not cirrhosis (OR 1·74, CI 0·87 – 3·49). Conversely, when using imaging, an association was demonstrated for cirrhosis (OR 2·26, CI 1·34 – 3·82) but not fibrosis (OR 1·69, CI 0·95 – 2·99). Concerning the types of singular infections, coinfection was 2·44 (95% CI 1·09 – 5·45) times more likely to cause any liver pathology than singular infection with HCV alone, and 2·33 (CI 0·46 – 11·86) times more likely than singular infection with a *Schistosoma* species, although this result was not significant.

**Table 2:** Subgroup analyses for any liver pathology. * 4 studies with unclear region (Kamal, 2001; Kamal, 2004; Houlihan, 2011 and el-Masry, 2006) removed from this subgroup analysis. † Sub-Saharan African region removed from this subgroup analysis due to insufficient studies in this group after removal of studies containing controls within the reference population. ‡ Combined hepatitis B and C subgroup removed from this subgroup analysis due to insufficient studies in this group after removal of studies containing controls within the reference population. § Two studies (Helal, 1998 and Hassan, 2003) focusing both on *S. mansoni* and *S. haematobium* removed from this subgroup analysis. ¶ Three studies (Gobert, 2015; Gunda, 2020; Houlihan, 2011) with unclear aim removed from this subgroup analysis. || Biomarker and epidemiology subgroups removed from this subgroup analysis due to insufficient studies in this group after removal of studies containing controls within the reference population. **One study (Houlihan, 2011) removed from this subgroup analysis due to only being a conference abstract. †† 6 studies with reference populations with mixed singular infections (Gunda, 2020; Allam, 2024; Kamal, 2000; Li, 2011; Iida, 1999 and Uchimura, 1997) as well as one study with a reference population with singular HBV infection (Houlihan, 2011) as well as 10 studies that included reference populations with health controls removed from this analysis.

| Subgroups | Studies | Number of participants | Number of co-infected individuals | Number of individuals with any liver pathology | OR (95% CI) | I<sup>2</sup> | Test of group differences |
| --- | --- | --- | --- | --- | --- | --- | --- |
| World Bank Region* |  |  |  |  |  |  |  |
| East Asia/Pacific | 7 | 2877 | 547 | 471 | 3.76 (1.74 – 8.14) | 65.47% | Qb (2) = 2.08, p= 0.35 |
| Middle East/ North Africa | 18 | 2086 | 763 | 854 | 2.07 (1.30 – 3.31) | 75.73% |  |
| Sub-Saharan Africa | 4 | 2710 | 140 | 318 | 2.00 (1.26 – 3.16) | 0.00% |  |
| World Bank Region subgroup with 10 studies including healthy controls removed<sup>†</sup> |  |  |  |  |  |  |  |
| East Asia/Pacific | 5 | 1243 | 206 | 281 | 3.25 (1.38 – 7.66) | 73.89% | Qb (1) = 1.23, p = 0.27 |
| Middle East/North Africa | 13 | 1305 | 570 | 570 | 1.81 (1.02 – 3.23) | 76.05% |  |
| Hepatitis species |  |  |  |  |  |  |  |
| B | 5 | 728 | 151 | 212 | 4.11 (1.46 – 11.59) | 77.65% | Qb (2) = 1.53, p= 0.47 |
| B and C | 6 | 2515 | 544 | 604 | 2.00 (1.09 – 3.69) | 44.29% |  |
| C | 22 | 4972 | 870 | 1056 | 2.78 (1.70 – 4.54) | 78.79% |  |
| Hepatitis species subgroup with 10 studies including healthy controls removed<sup>‡</sup> |  |  |  |  |  |  |  |
| B | 4 | 711 | 144 | 204 | 4.01 (1.19 – 13.54) | 84.31% | Qb (1) = 0.49, p = 0.48 |
| C | 17 | 4151 | 659 | 724 | 2.46 (1.57 – 4.58) | 81.63% |  |
| Schistosoma species<sup>§</sup> |  |  |  |  |  |  |  |
| Japonicum | 7 | 2877 | 547 | 471 | 3.76 (1.74 – 8.14) | 65.47% | Qb (1) = 0.42, p= 0.52 |
| Mansoni | 24 | 5124 | 945 | 1282 | 2.80 (1.78 – 4.40) | 77.11% |  |
| Schistosoma species subgroup with 10 studies including healthy controls removed |  |  |  |  |  |  |  |
| Japonicum | 5 | 1243 | 206 | 281 | 3.25 (1.38 – 7.66) | 73.89% | Qb (1) = 0.11, p = 0.74 |
| Mansoni | 17 | 3767 | 681 | 744 | 2.70 (1.41 – 5.16) | 83.72% |  |
| Study aim <sup>¶</sup> |  |  |  |  |  |  |  |
| Biomarker | 6 | 842 | 240 | 455 | 3.46 (1.14 – 10.49) | 70.40% | Qb (3) = 10.41, p= 0.02 |
| Epidemiology | 3 | 2151 | 113 | 157 | 1.06 (0.65 – 1.74) | 0.00% |  |
| Immune response | 5 | 508 | 226 | 183 | 11.41 (1.65 – 78.78) | 92.62% |  |
| Pathogenesis | 14 | 4075 | 879 | 883 | 2.38 (1.45 – 3.92) | 73.85% |  |
| Aim subgroup with 10 studies including healthy controls removed |  |  |  |  |  |  |  |
| Immune response | 4 | 347 | 171 | 114 | 17.37 (1.32 – 228.08) | 91.73% |  |
| Pathogenesis | 11 | 1907 | 469 | 591 | 2.17 (1.21 – 3.91) | 78.22% |  |
| Risk of bias** |  |  |  |  |  |  |  |
| Medium/Low | 18 | 4050 | 642 | 802 | 3.01 (1.84 – 4.92) | 68.38% | Qb (1) = 0.52, p= 0.47 |
| High | 14 | 3913 | 916 | 1034 | 2.30 (1.33 – 3.98) | 79.15% |  |
| Risk of bias subgroup with 10 studies including healthy controls removed |  |  |  |  |  |  |  |
| Medium/Low | 12 | 2936 | 456 | 425 | 3.71 (1.53 – 9.02) | 83.82% | Qb(1) = 1.55, p = 0.21 |
| High | 10 | 1866 | 447 | 582 | 1.89 (1.07 – 3.37) | 76.97% |  |
| Singular infections in reference population<sup>††</sup> |  |  |  |  |  |  |  |
| Hepatitis C only | 13 | 3033 | 502 | 549 | 2.44 (1.09 – 5.45) | 85.31 | Qb (1) = 0.00, p = = 0.96 |
| Schistosomiasis only | 3 | 269 | 127 | 100 | 2.33 (1.20 – 11.86) | 88.60% |  |

Only one study (3·30%) was classed as having low risk of bias^40^, seventeen studies (51·51%) had moderate risk of bias^32,33,48,49,51,52,55,60,62,34,35,38,39,41,43,44,47^ and fourteen (42·42%) had high risk of bias^14,31,57–59,61,36,37,42,45,46,53,54,56^ (Appendix 1 pp21-35). Studies with a high RoB showed weaker associations of coinfection with any liver pathology when compared to studies with a low or moderate RoB (Table 2). There was no difference in significant associations by RoB for the specific liver outcome of fibrosis (Table S15 Appendix 1 p47). Evidence of some publication bias was observed with smaller studies of positive effects potentially underrepresented and an underestimated overall pooled effect size (Figure S1 Appendix 1 p56). When removing studies with a high risk of bias, analyses for any liver pathology (OR 3·29, CI 1·95), fibrosis (OR 2·31, CI 1·25-4·25) and cirrhosis (OR 2·26, CI 1·36-5·06) remained robust (Figures S7, S8, S9 Appendix 1 pp48-50). This subgroup analysis could not be performed for the outcome HCC, as all but one study^44^ in this group had high RoB.

## Discussion

Helminth infections of *S. mansoni* , *S. japonicum* and *S. mekongi* as well as the viral infections of HBV and HCV cause significant long-term and potentially life-threatening liver diseases.^2,6,27,63^ We conducted a systematic review and meta-analysis of 33 studies, covering 8637 participants from seven countries to estimate the influence of coinfection on nonspecific liver disease, liver fibrosis, cirrhosis, and HCC. Coinfection of *S. mansoni* or *S. japonicum* with HBV or HCV was associated with a higher likelihood of all outcomes when compared to singularly or uninfected individuals.

We found that coinfected individuals were 2·75 times more likely to have any liver pathology than singularly or uninfected individuals. These associations emphasise the worsening of pathology mostly when compared to singular infection. Results remained robust when analyses were rerun using only studies with singularly infected individuals for the reference group. Hence, our findings might suggest an interaction between intestinal schistosomes and viral hepatitis that needs exploration in further studies to identify whether this is an additive or multiplicative effect, and how this effect influences the immune response.

The effect of coinfections was similar across varying severities of liver outcomes (ORs 1·81 to 2·22) of fibrosis, cirrhosis, and HCC when compared to singularly infected individuals. This finding may suggest that coinfections have an active interaction or modulating effect for pathogenesis as opposed to representing concurrent injury, particularly when comparing singular HCV infection and coinfection, such as mechanisms proposed of schistosomal infection prolonging viral hepatitis carrier states thus enabling viral-driven hepatocyte damage.^9,17,19^ However, the definition of fibrosis and cirrhosis used across published studies was heterogeneous and based on varied diagnostic scoring systems if a definition was provided at all. The lack of a designated scoring tool being used, especially in the cirrhosis studies, blurs the boundary between severe fibrosis and cirrhosis, potentially leading to misclassification. Additional research is needed to prospectively assess whether coinfection not only results in more likely hepatic disease, but also greater severity by providing consistent staging systems across studies.

We found that the influence of coinfection was moderated by diagnostic modality. Coinfection and liver fibrosis were only significantly correlated when biopsy was used and was insignificant when imaging was used. This could be due to early stages of fibrosis not resulting in changes to liver architecture that may be distinguishable on traditional ultrasounds, which is the presumed type of modality used in all six studies employing imaging as the diagnostic modality for fibrosis. Contrarily, coinfection was insignificantly positively associated with cirrhosis diagnosed by biopsies and significantly positively associated with cirrhosis diagnosed by imaging. Studies on cirrhosis using biopsy,^31,35,36,41–43,49,59^ all of which were ultrasound-guided, used a designated scoring tool to identify cirrhosis, such as the Ishak^64^ or METAVIR^65^ scores. None of the studies using ultrasound only,^33,34,54,55^ had a clear scoring system to produce a consistent definition of cirrhosis, which could introduce observer bias in the assessment to have falsely over- estimated the effect of imaging on the diagnosis of cirrhosis. These results highlight a need for studies that identify a shared scoring system as well as potential comparative research on biopsy versus imaging to find an accurate diagnostic modality which can be validated for pathologies related to schistosome and viral coinfection.

There were higher odds of any liver disease being present in the context of *S. japonicum* infection when compared to *S. mansoni*, suggesting as shown elsewhere,^66^ a more rapid progression of disease in *S. japonicum* infection. The more severe progression may be due to more rapid larval migration following entry into the human host, significantly increased egg production by female worms, smaller eggs resulting in more ectopic lesions, or faster and more severe activation of pro-inflammatory pathways within the liver.^67^ Yet, from this analysis it is unclear to what extent the seemingly higher likelihood of liver pathologies were influenced by *S. japonicum* compared to *S. mansoni* as opposed to HBV infection vs. HCV infection, as there were not enough studies to construct these sub-groups. Most of the studies on *S. japonicum* and HBV coinfection were in China where there have been large-scale control efforts for schistosomiasis morbidity management, and HBV vaccination. Consequently, it is unclear whether the pathology observed is only residual disease from a singular infection and coinfection was simply acquired after the disease development without any interactive effects actually caused by coinfection.

This review revealed limitations of the existing literature. While we did not exclude studies based on geography or language, there was a disproportionate number of studies from Egypt compared to other regions of the world. We were only able to include four studies from sub-Saharan Africa, seven from East Asia/Pacific in our meta-analysis, and none from South America, which is also a highly co-endemic setting.^2,66^ Future studies are needed on *S. mansoni* and HBV in sub-Saharan Africa where there is high prevalence of these infections.^2,40^ The majority of the articles included in our review did not report any adjusted data, and most studies had moderate to high risk of bias. Adjusting for viral load and chronicity of infection, as well as hepatitis vaccination status, age and other comorbidities should be investigated in future studies. Prospective cohort designs, such as the ongoing SchistoTrack Cohort,^68^ may lend future insights into these variables.

In conclusion, intestinal schistosome and viral hepatitis coinfection worsens hepatic outcomes beyond the effects observed from singular infections. There remains a large gap in the literature on the understanding of the pathogenesis of coinfection, and validation of a clinical diagnostic suitable for fibrosis from both schistosomal and viral hepatitis. While the importance of considering coinfections in the management and prevention of HBV morbidity and mortality has been recognised by the latest WHO guidelines on HBV, ^8^ coinfection with schistosomes has been omitted and there still exist no guidelines for schistosomiasis morbidity management. This review provides a strong argument for the need to consider coinfections in guidelines for schistosomiasis and HBV or HCV morbidity management, as well as to coordinate vaccination campaigns and mass drug administration to reduce the global burden of liver disease attributable to schistosomiasis and HBV or HCV.

## DECLARATIONS

## Data Availability

All data extracted and used for this meta-analysis will be available upon publication or are available upon request from the corresponding author. This study used openly available data published within the articles eligible for this meta-analysis.

## Acknowledgements

We thank the SchistoTrack Research Group for wider support on meta-analyses and feedback. All authors had full access to all the data in the study and had final responsibility for the decision to submit for publication.

## Author contributions

Resources and supervision: GFC. Conceptualization: GFC. Data curation: LK, HC. Pre-registration protocol, data extraction tool, and risk of bias tool: LK, GFC. Data validation: LK, LW, HC. Investigation and methodology: LK, LW, HC, GFC. Formal analysis and visualization: LK. Writing – original draft: LK, GFC. Writing – review and editing: LK, LW, HC, GFC. Funding acquisition: GFC.

## Open access statement

This research was funded in whole, or in part, by the UKRI EPSRC [EP/X021793/1]. For the purpose of Open Access, the author has applied a CC BY public copyright licence to any Author Accepted Manuscript version arising from this submission.

