## Appendix 1 for "Influence of intestinal schistosome and hepatitis B or C coinfection on hepatic disease: a systematic review and meta-analysis"

Title
Influence of intestinal schistosome and hepatitis B and C coinfection on hepatic disease: a systematic review and meta-analysis

Authors
Laura Kmentt MD, Lauren Wilburn PhD, Huike Cheng MSc, Goylette F. Chami PhD*

Affiliations
Big Data Institute, Nuffield Department of Population Health, University of Oxford, Oxford, UK (L Kmentt MD, L Wilburn PhD, Huike Cheng MSc, GF Chami PhD)

### Table S1: 2020 PRISMA Checklist

| Section and Topic | Item # | Checklist item | Location where item is reported |
|---|---|---|---|
| **TITLE** | | | |
| Title | 1 | Identify the report as a systematic review. | Pg 1 |
| **ABSTRACT** | | | |
| Abstract | 2 | See the PRISMA 2020 for Abstracts checklist. | Pg 1 |
| **INTRODUCTION** | | | |
| Rationale | 3 | Describe the rationale for the review in the context of existing knowledge. | Pg 5-7 |
| Objectives | 4 | Provide an explicit statement of the objective(s) or question(s) the review addresses. | Pg 7 |
| **METHODS** | | | |
| Eligibility criteria | 5 | Specify the inclusion and exclusion criteria for the review and how studies were grouped for the syntheses. | Methods, appendix 1 |
| Information sources | 6 | Specify all databases, registers, websites, organisations, reference lists and other sources searched or consulted to identify studies. Specify the date when each source was last searched or consulted. | Methods, appendix 1 |
| Search strategy | 7 | Present the full search strategies for all databases, registers and websites, including any filters and limits used. | Appendix 1 |
| Selection process | 8 | Specify the methods used to decide whether a study met the inclusion criteria of the review, including how many reviewers screened each record and each report retrieved, whether they worked independently, and if applicable, details of automation tools used in the process. | Methods |
| Data collection process | 9 | Specify the methods used to collect data from reports, including how many reviewers collected data from each report, whether they worked independently, any processes for obtaining or confirming data from study investigators, and if applicable, details of automation tools used in the process. | Methods |
| Data items | 10a | List and define all outcomes for which data were sought. Specify whether all results that were compatible with each outcome domain in each study were sought (e.g. for all measures, time points, analyses), and if not, the methods used to decide which results to collect. | Methods |
| | 10b | List and define all other variables for which data were sought (e.g. participant and intervention characteristics, funding sources). Describe any assumptions made about any missing or unclear information. | Methods |
| Study risk of bias assessment | 11 | Specify the methods used to assess risk of bias in the included studies, including details of the tool(s) used, how many reviewers assessed each study and whether they worked independently, and if applicable, details of automation tools used in the process. | Methods, appendix 1 |
| Effect measures | 12 | Specify for each outcome the effect measure(s) (e.g. risk ratio, mean difference) used in the synthesis or presentation of results. | Methods |
| Synthesis methods | 13a | Describe the processes used to decide which studies were eligible for each synthesis (e.g. tabulating the study intervention characteristics and comparing against the planned groups for each synthesis (item #5)). | Methods, appendix 1, appendix 2 |
| | 13b | Describe any methods required to prepare the data for presentation or synthesis, such as handling of missing summary statistics, or data conversions. | Methods |
| | 13c | Describe any methods used to tabulate or visually display results of individual studies and syntheses. | Methods |
| | 13d | Describe any methods used to synthesize results and provide a rationale for the choice(s). If meta-analysis was performed, describe the model(s), method(s) to identify the presence and extent of statistical heterogeneity, and software package(s) used. | Methods |
| | 13e | Describe any methods used to explore possible causes of heterogeneity among study results (e.g. subgroup analysis, meta-regression). | Methods |
| | 13f | Describe any sensitivity analyses conducted to assess robustness of the synthesized results. | Appendix 1 |
| Reporting bias assessment | 14 | Describe any methods used to assess risk of bias due to missing results in a synthesis (arising from reporting biases). | Appendix 1 |
| Certainty assessment | 15 | Describe any methods used to assess certainty (or confidence) in the body of evidence for an outcome. | Appendix 1 |
| **RESULTS** | | | |
| Study selection | 16a | Describe the results of the search and selection process, from the number of records identified in the search to the number of studies included in the review, ideally using a flow diagram. | Results, Appendix 1 |
| | 16b | Cite studies that might appear to meet the inclusion criteria, but which were excluded, and explain why they were excluded. | Appendix 1 |
| Study characteristics | 17 | Cite each included study and present its characteristics. | Results, Appendix 1 |
| Risk of bias in studies | 18 | Present assessments of risk of bias for each included study. | Appendix 1 |
| Results of individual studies | 19 | For all outcomes, present, for each study: (a) summary statistics for each group (where appropriate) and (b) an effect estimate and its precision (e.g. confidence/credible interval), ideally using structured tables or plots. | Results, Appendix 1 |
| Results of syntheses | 20a | For each synthesis, briefly summarise the characteristics and risk of bias among contributing studies. | Appendix 1 |
| | 20b | Present results of all statistical syntheses conducted. If meta-analysis was done, present for each the summary | Results, |

| Section and Topic | Item # | Checklist item | Location where item is reported |
|---|---|---|---|
| | | estimate and its precision (e.g. confidence/credible interval) and measures of statistical heterogeneity. If comparing groups, describe the direction of the effect. | Appendix 1 |
| | 20c | Present results of all investigations of possible causes of heterogeneity among study results. | Results |
| | 20d | Present results of all sensitivity analyses conducted to assess the robustness of the synthesized results. | Appendix 1 |
| Reporting biases | 21 | Present assessments of risk of bias due to missing results (arising from reporting biases) for each synthesis assessed. | Results |
| Certainty of evidence | 22 | Present assessments of certainty (or confidence) in the body of evidence for each outcome assessed. | Results |
| **DISCUSSION** | | | |
| Discussion | 23a | Provide a general interpretation of the results in the context of other evidence. | Discussion |
| | 23b | Discuss any limitations of the evidence included in the review. | Discussion |
| | 23c | Discuss any limitations of the review processes used. | Discussion |
| | 23d | Discuss implications of the results for practice, policy, and future research. | Discussion, Research in context |
| **OTHER INFORMATION** | | | |
| Registration and protocol | 24a | Provide registration information for the review, including register name and registration number, or state that the review was not registered. | Abstract, Methods |
| | 24b | Indicate where the review protocol can be accessed, or state that a protocol was not prepared. | Abstract, Methods |
| | 24c | Describe and explain any amendments to information provided at registration or in the protocol. | Methods |
| Support | 25 | Describe sources of financial or non-financial support for the review, and the role of the funders or sponsors in the review. | Abstract, Funding statement |
| Competing interests | 26 | Declare any competing interests of review authors. | Declarations |
| Availability of data, code and other materials | 27 | Report which of the following are publicly available and where they can be found: template data collection forms; data extracted from included studies; data used for all analyses; analytic code; any other materials used in the review. | Supplement |

### Text S1: Search strategy

The search string was piloted on the five databases Cochrane Central Register of Controlled Trials (1996-), Medline including PubMed (1946-), Embase (1974-), Global Health (1973-) and Global Index Medicus (1901-) on 3rd July 2023 using a predefined search string. The updated and final search was carried out on 7th July on the same five databases and updated on 23rd January 2025. We did not place any restrictions on dates or language. Retrieved references were exported to the Endnote reference manager for initial de-duplication, and then to Covidence for further screening.

#### Table S2: Summary of hits on searches across the five databases

| Database | Interface | Coverage | Hits 3.7.23 | Hits 7.7.23 | Hits 23.1.25 |
|---|---|---|---|---|---|
| Medline | Ovid SP | | 405 | 648 | 17 |
| Embase | Ovid SP | | 799 | 1335 | 69 |
| Global Health | Ovid SP | | 333 | 519 | 18 |
| Global Index Medicus | http:// pesquisa.bv salud.org/gim/ | | 103 | 137 | 250 |
| Cochrane Central Register of Controlled Trials | Cochrane Library, Wiley | | 12 | 23 | 0 |
| Total | | | 1652 | 2662 | 254 |
| Duplicates removed in Covidence | | | | 1011 | 79 |
| **Final total** | | | | 1651 | **333** |

#### Table S3: Summary of Medline search terms

| # | Search terms | Results 7.7.23 | Results 23.1.25 |
|---|---|---|---|
| 1 | (schistosom* OR bilharzia* OR "snail fever" OR "mansoni" OR "japonicum" OR "mekongi") | 40796 | 1377 |
| 2 | ("hepatitis B" OR HBV OR "hepatitis C" or HCV or "hepatitis B/C").mp | 198091 | 8752 |
| 3 | (liver OR hepat* OR cirrho*).mp | 1582810 | 93120 |
| 4 | 1 AND 2 AND 3 | 648 | 17 |
| 5 | 1 AND 2 | 658 | 17 |
| 6 | 1 AND 3 | 7326 | 234 |
| 7 | 2 AND 3 | 194658 | 8450 |

[mp=title, book title, abstract, original title, name of substance word, subject heading word, floating sub-heading word, keyword heading word, organism supplementary concept word, protocol supplementary concept word, rare disease supplementary concept word, unique identifier, synonyms, population supplementary concept word, anatomy supplementary concept word]

#### Table S4: Summary of Embase search terms

| # | Search terms | Results 7.7.23 | Results 23.1.25 |
|---|---|---|---|
| 1 | (schistosom* OR bilharzia* OR "snail fever" OR "mansoni" OR "japonicum" OR "mekongi") | 43762 | 1278 |
| 2 | ("hepatitis B" OR HBV OR "hepatitis C" or HCV or "hepatitis B/C").mp | 339181 | 14757 |
| 3 | (liver OR hepat* OR cirrho*).mp | 2220530 | 143860 |
| 4 | 1 AND 2 AND 3 | 1335 | 69 |
| 5 | 1 AND 2 | 1351 | 70 |
| 6 | 1 AND 3 | 8687 | 355 |
| 7 | 2 AND 3 | 333414 | 14664 |

[mp=title, abstract, heading word, drug trade name, original title, device manufacturer, drug manufacturer, device trade name, keyword heading word, floating subheading word, candidate term word]

#### Table S5: Summary of Global Health search terms

| # | Search terms | Results 7.7.23 | Results 23.1.25 |
|---|---|---|---|
| 1 | (schistosom* OR bilharzia* OR "snail fever" OR "mansoni" OR "japonicum" OR "mekongi") | 37611 | 1736 |
| 2 | ("hepatitis B" OR HBV OR "hepatitis C" or HCV or "hepatitis B/C").mp | 75996 | 4468 |
| 3 | (liver OR hepat* OR cirrho*).mp | 290988 | 31224 |
| 4 | 1 AND 2 AND 3 | 519 | 18 |
| 5 | 1 AND 2 | 523 | 18 |

| | | | |
|---|---|---|---|
| 6 | 1 AND 3 | 6601 | 240 |
| 7 | 2 AND 3 | 75445 | 4421 |

[mp = abstract, title, original title, heading words, cabicodes words]

#### Table S6: Summary of Global Index Medicus search terms

| # | Search terms | Results 7.7.23 | Results 23.1.25 |
|---|---|---|---|
| 1 | (schistosom* OR bilharzia* OR "snail fever" OR “mansoni” OR “japonicum” OR “mekongi”) | 8160 | 173 |
| 2 | (“hepatitis B” OR HBV OR “hepatitis C” or HCV or “hepatitis B/C”).mp | 22862 | 276 |
| 3 | (liver OR hepat* OR cirrho*).mp | 129203 | 7853 |
| 4 | 1 AND 2 AND 3 | 137 | 150 |
| 5 | 1 AND 2 | 137 | 151 |
| 6 | 1 AND 3 | 2522 | 32 |
| 7 | 2 AND 3 | 22139 | 263 |

[tw: title, abstract, subject]

#### Table S7: Summary of Cochrane Central Register of Controlled Trials search terms

| # | Search terms | Results 7.7.23 | Results 23.1.25 |
|---|---|---|---|
| 1 | (schistosom* OR bilharzia* OR "snail fever" OR “mansoni” OR “japonicum” OR “mekongi”) | 798 | 866 |
| 2 | (“hepatitis B” OR HBV OR “hepatitis C” or HCV or “hepatitis B/C”).mp | 19644 | 20616 |
| 3 | (liver OR hepat* OR cirrho*).mp | 88590 | 110125 |
| 4 | 1 AND 2 AND 3 | 23 | 23 |
| 5 | 1 AND 2 | 24 | 24 |
| 6 | 1 AND 3 | 131 | 143 |
| 7 | 2 AND 3 | 84 | 20041 |

[title, abstract, keyword]

#### Table S8: PECO inclusion criteria

| | | |
|---|---|---|
| **P** | Participants | • Any age or sex<br>• Living in endemic areas for *S. mansoni, S. japonicum, S. mekongi* |
| **E** | Exposure | Current or past intestinal *Schistosoma* and hepatitis B and/or C co-infection:<br>• Binary variable (yes/no).<br>• Eligible intestinal *Schistosoma* species (*S. mansoni, S. japonicum, S. mekongi*).<br>• Hepatitis species are B and/or C.<br>• Schistosomiasis infection diagnosed using antigenic, antibody, microscopic methods.<br>• If imaging modality is used to diagnose schisosomiasis infection (e.g. ultrasound appearances suggestive or schistosomiasis) this must have also been confirmed using any of the above laboratory testing modalities.<br>• Past infection must have been diagnosed using valid diagnostic methods.<br>• Hepatitis infection diagnosed using antigen, antibody or viral load diagnostic tests. |
| **C** | Comparator | Individuals infected with only one of the pathogens of interest (*S. mansoni, S. japonicum, S. mekongi,* hepatitis B virus, hepatitis C virus) or no infection. |
| **O** | Outcome | The outcomes of interest are the following four liver pathologies: hepatomegaly, fibrosis, cirrhosis and hepatocellular carcinoma. These were coded as binary variables (yes/no).<br>• Author-defined hepatomegaly diagnosed using imaging modality or clinical examination with reference population given.<br>• Author-defined liver fibrosis diagnosed using imaging (MRI, CT, PET, USS), biomarkers (where authors have explicitly stated that these are for use as assessment of fibrosis and not as proxy for liver function) or biopsy.<br>• Author-defined liver cirrhosis diagnosed using imaging (MRI, CT, PET, USS), biomarkers (where authors have explicitly stated that these are for use as assessment of cirrhosis and not as proxy for liver function) or biopsy.<br>• Author-defined hepatocellular carcinoma diagnosed using imaging (MRI, CT, PET, USS), biopsy or biomarkers (tumour markers) in conjunction with either imaging or biopsy. |

### Table S9: Full list of excluded studies with reasons for exclusion

| Title | Study | Reason for exclusion |
|---|---|---|
| HBV serological markers versus clinico-biochmical, and histopathological evidence of activity in the evaluation of cirrhotic patients group A Child's Classification with hematemesis and/or melena | A 1990 | Full text article could not be obtained |
| Hepatitis B virus markers in schistosomiasis patients in a village menoufia governorate, Egypt, and their response to praziquantel | A 1993 | Full text article could not be obtained |
| Gastric emptying time in bilharzial patients with and without hepatitis-C virus infection | A 1994 | Full text article could not be obtained |
| Significance of iomantibody to hepatitis C virus in Egyptian patients with chronic hepatitis C | A 1994 | Full text article could not be obtained |
| Role of bilharziasis in the persistence of hepatitis B antigenaemia | A 1994 | Full text article could not be obtained |
| Association of Hepatitis C Virus and endemic Hepatosplenomegaly | A 1995 | Full text article could not be obtained |
| Hepatitis C virus [HCV] markers in patients with schistosomiasis a study in an Egyptian village | A 1995 | Full text article could not be obtained |
| Assessment of liver dysfunction in silica exposed workers | A 1998 | Full text article could not be obtained |
| Seroprevalence and risk factors for hepatitis B and C virus infection in Damietta Governorate, Egypt | A 2014 | Wrong outcomes |
| The role of BCL9 genetic variation as a biomarker for hepatitis C-related hepatocellular carcinoma in Egyptian patients | Abbas 2022 | Insufficient information to calculate outcome effect sizes |
| Serum Visfatin in patients with chronic hepatitis C | Abd-El-Fatah 2011 | Wrong outcomes |
| Dysregulation of blood lymphocyte subsets and natural killer cells in schistosomal liver cirrhosis and hepatocellular carcinoma | AbdAlFattah 2003 | Wrong outcomes |
| Diagnostic and prognostic value of direct and indirect non-invasive bio-markers versus liver biopsy to stage-hepatic fibrosis in patients with isolated chronic HCV and co-infected with schistosomiasis | Abdel-Aziz 2012 | Insufficient information to calculate outcome effect sizes |
| Clinical Benefits of Biochemical Markers of Fibrosis in Egyptian Children With Chronic Liver Diseases | Abdel-Ghaffar 2010 | Wrong outcomes |
| Causes of minimal hepatic periportal fibrosis present in Egypt | Abdel-Kader 1997 | No reference population given |
| Coinfection with hepatitis C virus and schistosomiasis: fibrosis and treatment response | Abdel-Rahman 2013 | Insufficient information to calculate outcome effect sizes |
| Schistosomiasis mansoni in an Egyptian village in the Nile delta | Abdel-Wahab 1980 | Wrong outcomes |
| Epidemiology of hepatocellular carcinoma in lower Egypt, Mansoura Gastroenterology Center | Abdel-Wahab 2007 | Non-eligible diagnostic method used for schistosomiasis |
| Liver Disease Outcomes after Sustained Virological Response in Patients with Chronic Hepatitis C Infection Treated with Generic Direct-Acting Antivirals | AbdEl-Wahab 2022 | Wrong outcomes |
| The role of intrahepaticT-Cell in immunopathogenesis of the liver cirrhosis in chronic hepatitis C with and without Schistosomiasis | Abdelaal 2013 | Full text article could not be obtained |
| <The> pathogenesis of cytokines in preportal fibrosis of human infected with schistosomiasis and viral hepatitis | AbdelAtty 2005 | Wrong outcomes |
| Lack of association between schistosomiasis and hepatitis B virus infection in Gezira-Managil area, Sudan | AbdellaEltoum 1991 | Insufficient information to calculate outcome effect sizes |
| Study of portal and systemic levels of nitric oxide, endothelin-1 and procollagen III peptide in chronic liver disease in Egypt | AbdElMoety 2010 | Full text article could not be obtained |
| Histocompatibility antigens in relation to hepatic Schistosomiasis | AbdElMoety 2010 | Full text article could not be obtained |
| PREVALENCE AND ASSOCIATED RISK FACTORS OF HEPATITIS C IN OLDER EGYPTIAN PATIENTS | Abou-Raya 2022 | Full text article could not be obtained |
| Coinfection of Schistosoma Species with Hepatitis B or Hepatitis C Viruses | Abruzzi 2016 | Review/editorial |
| Study of serum levels of interferon-gamma [IFN-Gamma] and human tumour T necrosis factor-Alpha [TNF-Alpha] in chronic hepatitis C with or without Bilharziasis | Adel 1999 | Full text article could not be obtained |
| Study of interleukin-6 and interleukin-8 in Egyptian patients with chronic liver disease due to hepatitis C and bilharziasis | AdelAhmed 2000 | Full text article could not be obtained |
| Androgen profiles among Egyptian adults considering liver status | Aguilar 2008 | Wrong outcomes |
| Alpha-Glutathione S-transferase and serum aminotransferases in schistosomiasis mansoni patients with or without hepatitis C virus | Ahmed 2008 | No reference population given |
| Hepatocellular carcinoma in patients with parasitic liver disease--Japanese schistosomiasis and hepatocellular carcinoma | Akahane 2001 | Full text article could not be obtained |
| Localization of schistosoma mansoni antigen confirms the frequent coexistence of hepatic schistosomiasis with chronic viral hepatitis C. An immunohistochemical study | AklMahmoud 2009 | Full text article could not be obtained |
| Hepatitis B virus (HBV) markers among patients with chronic liver disease in Kuwait | Al-Nakib 1982 | Wrong outcomes |

| | | |
|---|---|---|
| Patterns of chronic liver disease in Kuwait with special reference to localisation of hepatitis B surface antigen | AlAdnani 1984 | Wrong outcomes |
| Alterações duodenais na hipertensão portal da esquistossomose mansônica | Almeida 2015 | Wrong outcomes |
| Digestive system malignancies in the eastern province of Saudi Arabia: an analysis of 158 patients | alQuorain 1988 | Full text article could not be obtained |
| HCV and associated concomitant infections at Sharkia Governorate Egypt | Amal 2004 | Full text article could not be obtained |
| Advanced hepatic schistosomiasis and chronic viral hepatitis | Andrade 1977 | Full text article could not be obtained |
| Active chronic hepatitis and decompensated schistosomiasis | Andrade 1978 | Full text article could not be obtained |
| Pathology of human schistosomiasis | Andrade 1987 | Review/editorial |
| Chronic hepatitis B and liver schistosomiasis: A deleterious association | Andrade 2014 | Insufficient information to calculate outcome effect sizes |
| Chronic liver disease in the Alexandria governorate, Egypt: Contribution of schistosomiasis and hepatitis virus infections | Angelico 1997 | Insufficient information to calculate outcome effect sizes |
| Hepatitis B and C virus markers among patients with hepatosplenic mansonic schistosomiasis | Aquino 2000 | Wrong outcomes |
| Epidemiologicalaaspects of hepatocellular carcioma in a referral center of minnas gerais, Brazil | Arquivosde 2013 | No reference population given |
| Cytokines and immunoglobulin-E in certain parasitic diseases | Atef 2006 | Full text article could not be obtained |
| Impact of hepatitis C virus/schistosoma mansoni coinfection on the circulating levels of HCV-NS4 protein and extracellular-matrix deposition in patients with different hepatic fibrosis stages | Attallah 2016 | Insufficient information to calculate outcome effect sizes |
| Levels of Schistosoma mansoni circulating antigen in chronic hepatitis c patients with different stages of liver fibrosis | Attallah 2016 | Wrong outcomes |
| Prevalence, impact and risk factors of hepatitis C infection | Azza 1993 | Wrong outcomes |
| Natural history of hepatitis C virus among apparently normal schoolchildren: follow up after 7 years | Azza 2004 | Review/editorial |
| Evaluation of different health hazards among pharmacists and their assistants at Cairo university hospitals [CUH] | B 2000 | Full text article could not be obtained |
| Risk factors for hepatocellular carcinoma in Egypt: The role of hepatitis-B viral infection and schistosomiasis | Badawi 1999 | Non-eligible diagnostic method used for schistosomiasis |
| Hepatic expression of nitric oxide isoforms and serum nitrites/nitrates in chronic hepatitis C and schistosomal liver disease | Badawy 2011 | Full text article could not be obtained |
| Schistosoma mansoni soluble egg antigens enhance HCV replication in mammalian cells | Bahgat 2010 | Wrong outcomes |
| Chronic hepatitis B antigenaemia in patients with hepatosplenic schistosomiasis | Bassily 1979 | Full text article could not be obtained |
| Chronic hepatitis B patients with schistosomiasis mansoni | Bassily 1983 | Full text article could not be obtained |
| Immunogenicity of hepatitis B vaccine in patients infected with Schistosoma mansoni | Bassily 1987 | Wrong outcomes |
| Hepatitis C virus infection and hepatosplenic schistosomiasis | Bassily 1992 | Review/editorial |
| Hepatitis viruses, schistosomal infection and liver cancer in Egypt | Bedwani 1996 | Review/editorial |
| Intensity of Schistosoma mansoni, hepatitis B, age, and sex predict levels of hepatic periportal thickening/fibrosis (PPT/F): A large-scale community-based study in Ethiopia | Berhe 2007 | Wrong outcomes |
| Reversibility of schistosomal periportal thickening/fibrosis after praziquantel therapy: A twenty-six month follow-up study in Ethiopia | Berhe 2009 | Full text article could not be obtained |
| Population-based differences in Schistosoma mansoni- and hepatitis C-induced disease | Blanton 2002 | Insufficient information to calculate outcome effect sizes |
| High prevalence of anti-HEV IgG is not associated with HIV infection in Rakai, Uganda | Boon 2016 | Ineligible hepatitis species |
| The association of chronic hepatitis B infection with chronic schistosomiasis mansoni in Kenya | Bosch 1987 | Wrong outcomes |
| Preliminary investigation on serum markers of hepatitis B virus in patients with Schistosomiasis japonica | Cai 1985 | Wrong outcomes |
| Hepatitis B and schistosomiasis: interaction or no interaction? | Chen 1993 | Full text article could not be obtained |
| Portopulmonary hypertension in cirrhotic patients: Prevalence, clinical features and risk factors | Chen 2013 | Wrong outcomes |
| Diagnostic characteristics and hepatic histopathology in 115 patients with liver injury of unknown reasons | Chen 2014 | Wrong outcomes |
| Prevention and treatment of intestinal bacterial translocation in patients with dycompensated cirrhosis by Tongfu method and its mechanism | ChiCtr 2018 | Insufficient information to calculate outcome effect sizes |
| PREVALENCE AND CLINICAL SIGNIFICANCE OF SCHISTOSOMIASIS-CHRONIC HEPATITIS B VIRUS CO-INFECTION IN ZAMBIA | Chisenga 2017 | Full text article could not be obtained |
| Duplex Doppler ultrasound of hepatic Schistosomiasis japonica: A study of 47 patients | Chou 2003 | Insufficient information to calculate outcome effect sizes |

| | | |
|---|---|---|
| Chronic splenomegaly in Nairobi, Kenya. II. Portal hypertension | Cock 1987 | Wrong outcomes |
| Prognosis of schistosomiasis mansoni patients infected with hepatitis B virus | Conceicao 1998 | Wrong outcomes |
| Hepatitis B and Schistosoma co-infection in a non-endemic area | Cuenca-Gomez 2016 | Non-endemic setting |
| Investigate of the etiology and prevention status of liver cirrhosis | Dai 2023 | Wrong outcomes |
| Hepatitis B virus infection among immunocompromised patients in Egypt | Darwish 1990 | Wrong outcomes |
| Hepatitis C and cirrhotic liver disease in the Nile delta of Egypt: A community-based study | Darwish 2001 | Non-eligible diagnostic method used for schistosomiasis |
| Morbidity associated with schistosomiasis before and after treatment in young children in Rusinga Island, Western Kenya | Davis 2015 | Wrong outcomes |
| Duodenal changes in portal hypertension schistosomiasis | DeAlmeida 2015 | Wrong outcomes |
| Portal hypertension in Nairobi, Kenya | deCock 1983 | Wrong outcomes |
| Correlation of biological serum markers with the degree of hepatic fibrosis and necroinflammatory activity in hepatitis C and schistosomiasis patients | deMorais 2010 | Insufficient information to calculate outcome effect sizes |
| Treatment of hepatitis C virus genotype 4 with peginterferon alfa-2a: impact of bilharziasis and fibrosis stage | Derbala 2006 | Wrong outcomes |
| Anti-HCV IgM as predictor of response to interferon therapy in schistosomal patients with chronic hepatitis C [1] | Derbala 2006 | Review/editorial |
| Hepatitis C genotype 4 with normal transaminases and schistosomiasis co-infection: Histological changes, response rate, late relapse, and hematological adverse effect | Derbala 2010 | Full text article could not be obtained |
| Hepatitis C virus genotype 4 with normal transaminases: Histological changes, schistosomiasis and response to treatment | Derbala 2011 | Insufficient information to calculate outcome effect sizes |
| Aspartate transaminase to platelet ratio index (APRI) in HCV and Schistosomiasis coinfection | Derbala 2016 | Full text article could not be obtained |
| Variants of CTGF are associated with hepatic fibrosis in Chinese, Sudanese, and Brazilians infected with Schistosomes | Dessein 2009 | Wrong outcomes |
| Relationship between advanced schistosomiasis and HBV infection | Du 2013 | Wrong outcomes |
| The Prevalence, Predictors, and In-Hospital Mortality of Hepatic Encephalopathy in Patients with Liver Cirrhosis Admitted at St. Dominic Hospital in Akwatia, Ghana | Duah 2020 | Wrong outcomes |
| Circulating markers of oxidative stress and liver fibrosis in Sudanese subjects at risk of schistosomiasis and hepatitis | Eboumbou 2005 | Non-eligible diagnostic method used for schistosomiasis |
| Role of liver biopsy in the management of liver diseases following the end of interferon era: Experience of a tertiary referral center | Ehsan 2019 | Full text article could not be obtained |
| Hepatitis C virus and schistosomiasis as a causative factor for hTERT amplification in hepatocellular carcinoma | Eid 2016 | Wrong outcomes |
| Soluble egg antigen of Schistosoma Haematobium induces HCV replication in PBMC from patients with chronic HCV infection | El-Awady 2006 | Wrong Schistosoma species |
| Circulating intercellular adhesion molecule-1 in endemic chronic liver diseases | El-Gindy 1997 | Wrong outcomes |
| Delta virus versus HBsAg in chronic active hepatitis and their relation to clinical, laboratory, and morbidity findings in bilharzial and non-bilharzial patients | el-Hawey 1993 | Wrong outcomes |
| Chronic liver disease and hepatitis C virus in Egyptian patients | El-Medany 1999 | Non-eligible diagnostic method used for schistosomiasis |
| Role of CCR5DELTA32 mutation in protecting patients with Schistosoma mansoni infection against hepatitis C viral infection or progression | El-Moamly 2013 | Wrong outcomes |
| Prevalence of hepatitis-C antibody seropositivity in healthy Egyptian children and four high risk groups | El-Nanaway 1995 | Wrong outcomes |
| The prevalence of hepatitis B and C infections among immigrants to a newly reclaimed area endemic for Schistosoma mansoni in Sinai, Egypt | El-Sayed 1997 | Wrong outcomes |
| 99mTechnetium-macroaggregated albumin perfusion lung scan versus contrast enhanced echocardiography in the diagnosis of the hepatopulmonary syndrome in children with chronic liver disease | El-Shabrawi 2010 | Wrong outcomes |
| Non-invasive markers and predictors of severity of hepatic fibrosis in HCV patients at Sharkia Governorate, Egypt | El-Shorbagy 2004 | Full text article could not be obtained |
| A prospective randomized trial comparing medical versus endoscopic treatment in prevention of variceal rebleeding in patients with schistosomal hepatic fibrosis co-infected with HCV | El-Tahawy 2002 | Full text article could not be obtained |
| Human Schistosomiasis mansoni associated with hepatocellular carcinoma in Egypt: current perspective | El-Tonsy 2016 | Insufficient information to calculate outcome effect sizes |
| Does schistosomiasis play a role in the high sero prevalence of HCV antibody among Egyptians? | el-Zayadi 1997 | Wrong outcomes |
| Assessment of liver function by the MEGX test in patients with schistosomiasis and cirrhosis | ElDesoky 1999 | Wrong outcomes |
| Egy-score can predict portal hypertension in chronic hepatitis C with good accuracy | Elghamry 2016 | Full text article could not be obtained |
| High prevalence of hepatitis C virus among urban and rural population groups in Egypt | elGohary 1995 | Wrong outcomes |
| Insulin resistance: A major predictor of significant liver fibrosis in Egyptian patients with genotype 4 chronic hepatitis C | ElRay 2010 | Full text article could not be obtained |

| | | |
|---|---|---|
| Demographic, epidemiological, clinical, severity, and treatment response characteristics of hepatitis b Egyptian patients | Elsabaawy 2018 | Full text article could not be obtained |
| Possible contribution of serum activin A and IGF-1 in the development of hepatocellular carcinoma in Egyptian patients suffering from combined hepatitis C virus infection and hepatic schistosomiasis | Elsammak 2006 | Wrong outcomes |
| IL-4 and reactive oxygen species are elevated in Egyptian patients affected with schistosomal liver disease | Elsammak 2008 | Wrong outcomes |
| HCV/schistosomiasis coinfection: Impact on fibrosis and response to pegylated interferon and ribavirin therapy | Elsharkawy 2012 | Full text article could not be obtained |
| Prognostic significance of hepatic veins waveforms study by doppler ultrasonography in cirrhotic patients with portal hypertension | Eman 2000 | Full text article could not be obtained |
| Hepatitis C infection among Egyptian blood donors in the eastern Saudi Arabia with / without past history of schistosomiasis | Fathalla 1994 | Wrong outcomes |
| Valsartan plus sclerotherapy compared with sclerotherapy alone in cirrhotic patients after variceal bleeding : clinical and haemodynamic randomized trial | Fayza 2005 | Full text article could not be obtained |
| IS SCHISTOSOMIASIS A RISK FACTOR OF CHOLANGIOCARCINOMA? | Forlemu 2023 | Full text article could not be obtained |
| Hepatic schistosomiasis | Fung 2009 | Review/editorial |
| Alpha one antitrypsin relation tochronic viral hepatitis c and b and h.c.c. in chronic liver disease | Galal 2006 | Full text article could not be obtained |
| Histocompatibility antigens in bilharzial hepatic fibrosis | Gamal 1986 | Full text article could not be obtained |
| Outcomes in 161 patients with decompensated hepatitis B: A single center study | Gao 2010 | Full text article could not be obtained |
| Assessment of health status of workers in a major waste dumpsite with special emphasis on liver disease | Gehad 2003 | Full text article could not be obtained |
| Hepatitis B vaccination in children infected with Schistosoma mansoni: Correlation with ultrasonographic data | Ghaffar 1990 | Wrong outcomes |
| Comparison of hepatic and splenic stiffness in chronic hepatitis C infection with and without schistosomal infection; Correlation with hepatic histopathological changes | Gohary 2020 | No reference population given |
| Liver biopsy findings in the acquired immunodeficiency syndrome | González 1988 | Full text article could not be obtained |
| Diagnosis of coinfection by schistosomiasis and viral hepatitis B or C using 1H NMR-based metabonomics | Gouveia 2017 | Non-eligible diagnostic method used for schistosomiasis |
| Positive false reaction of HBsAg (by RPHA) in patients with schistosomiasis and its mechanism | Gui 1986 | Full text article could not be obtained |
| The detection and significance of HBV-DNA in serum and liver of advanced schistosomiasis patients | Gui 1994 | Full text article could not be obtained |
| A clinical and pathological study on patients with splenomegalic advanced schistosomiasis | Gui 1997 | Full text article could not be obtained |
| [Hepatitis B antigens systems in schistosomiasis mansoni] | Guimaraes 1981 | Full text article could not be obtained |
| Incidence and meaning of hepatitis B antigenic systems in schistosomiasis | Guimaraes 1981 | Full text article could not be obtained |
| Hepatitis C viraemia and cortisol level in thalassaemic children along with other viraemia risk factors | H 2000 | Full text article could not be obtained |
| Effect of schistosomiasis and hepatitis on liver disease | Halim 1999 | Insufficient information to calculate outcome effect sizes |
| HBSAg antigenaemia in bilharzial patients | Hamadto 1989 | Wrong outcomes |
| Study on some hepatic functions and prevalence of hepatitis B surface antigenaemiain Egyptian children with schistosomal hepatic fibrosis | Hammad 1990 | Wrong outcomes |
| <A> cross sectional study of hepatitis B, C, some trace elements, heavy metals, aflatoxin B1 and schistosomiasis in a rural population, Egypt | HananAli 2005 | Wrong outcomes |
| The role of hepatitis C in hepatocellular carcinoma: A case control study among egyptian patients | Hassan 2001 | Insufficient information to calculate outcome effect sizes |
| Evaluation of nitric oxide (NO) levels in hepatitis C virus (HCV) infection: Relationship to schistosomiasis and liver cirrhosis among Egyptian patients | Hassan 2002 | Wrong outcomes |
| Serum hepatitis B virus (HBV) markers and aminoterminal peptide of type III procollagen (PIIINP) levels in patients with hepatosplenic schistosomiasis in Egypt | Hassanein 1989 | Full text article could not be obtained |
| CD30 in patients with HVC-related cirrhosis and HCC - Is it a marker of disease activity? | Hassoba 2004 | Non-eligible diagnostic method used for schistosomiasis |
| Evolution of living donor liver transplantation in Egypt | Hatem 2005 | Wrong outcomes |
| Schistosomiasis | Hatz 2009 | Full text article could not be obtained |
| Laparoscopic diagnosis and clinical course of chronic schistosomiasis japonica | Hayashi 2000 | Non-eligible diagnostic method used for schistosomiasis |
| Association of hepatitis B infection with schistosomiasis in children | Hesham 1990 | No reference population given |
| Granulocyte macrophage colony stimulating factor: role in induction of monocyte CD14 receptor and adhesion molecules expression in chronic liver diseases | Hesham 2003 | Full text article could not be obtained |

| | | |
|---|---|---|
| The impact of schistosomiasis among rural populations in Liberia | Holzer 1983 | Wrong outcomes |
| <The> frequency of Anti-HCV among Egyptian rural school children: Its relation to schistosomiasis | Hosny 1995 | Full text article could not be obtained |
| Study on the clinical usefulness of the serum fibrosis index to diagnose hepatic fibrosis in patients with schistosomiasis | Hu 2014 | Full text article could not be obtained |
| A comparative study of intrahepatic cholangiocarcinoma and hepatocellular carcinoma with reference to clinical features and prognosis | Hu 2019 | Wrong outcomes |
| An analysis of 240 newly recurrent advanced schistosomiasis cases in Shashi District of Hubei Province in 2004-2009 | Huang 2011 | Wrong outcomes |
| The efficacy and safety of entecavir in patients with advanced schistosomiasis co-infected with hepatitis B virus | Huang 2013 | No reference population given |
| Analysis of factors affecting the occurrence of advanced schistosomiasis japonica in Poyang Lake area | Huang 2021 | Full text article could not be obtained |
| Parenteral antischistosomal therapy: A potential risk factor for hepatitis B infection | Hyams 1987 | Insufficient information to calculate outcome effect sizes |
| A case-control study on liver cancer with special emphasis on the possible aetiological role of schistosomiasis | Inaba 1984 | No reference population given |
| Liver cancer in an endemic area of schistosomiasis japonica in Yamanashi Prefecture, Japan | Inaba 1988 | Full text article could not be obtained |
| Hepatitis B virus markers in patients with schistosomiasis, liver cirrhosis and hepatocellular carcinoma in Khartoum, Sudan | Itoshima 1989 | Wrong outcomes |
| Quantifying quality of life and disability of patients with advanced schistosomiasis japonica | Jia 2011 | Insufficient information to calculate outcome effect sizes |
| Clinicopathological study on correlation between advanced schistosomiasis and hepatitis B | Jiang 1988 | No reference population given |
| Cases of advanced schistosomiasis associated with positive HBsAg | Jin 1986 | Full text article could not be obtained |
| Antioxidant status and lipid peroxidation activity in evaluating hepatocellular damage in children | K 2009 | Wrong outcomes |
| Prevalence of hepatitis B surface antigen in serum and ascitic fluid of patients with chronic liver diseases | Kabil 1990 | Full text article could not be obtained |
| The true situation of liver diseases in Egypt with the beginning of declension of virus B hepatitis | Kabil 1990 | Full text article could not be obtained |
| Study of HBV markers and HLA typing in chronic liver diseases (CLD) | Kabil 1991 | Wrong outcomes |
| Interferon therapy in patients with chronic hepatitis C and schistosomiasis | Kamal 2000 | Review/editorial |
| Progression of fibrosis in hepatitis C with and without schistosomiasis: Correlation with serum markers of fibrosis | Kamal 2006 | Insufficient information to calculate outcome effect sizes |
| The epidemiology of Schistosoma mansoni, hepatitis B and hepatitis C infection in Egypt | Kamel 1994 | Insufficient information to calculate outcome effect sizes |
| Assessment of splenic functions in patients with hepato-splenic schistosomiasis using non-invasive techniques | Kamel 1999 | Wrong outcomes |
| P selectin and T cell profiles provide verification to understand the pathogenesis of liver cirrhosis in HCV and Schistosoma mansoni infections | Kamel 2014 | Wrong outcomes |
| P Selectins and immunological profiles in HCV and Schistosoma mansoni induced chronic liver disease | Kamel 2014 | Wrong outcomes |
| Plasma and tissue fibronectin and serum procollagen iii peptide in chronic liver disease patients as reliable biomarkers for hepatic fibrogenesis | Kamel 2015 | Full text article could not be obtained |
| Serum soluble transferrin receptors in relation to iron status in Egyptian chronic viral hepatitis C patients with and without schistosomal hepatic fibrosis | Kandil 2011 | Full text article could not be obtained |
| Hepatosplenic morbidity in schistosomiasis japonica: Evaluation with doppler sonography | Kardorff 1999 | Wrong outcomes |
| Immune receptors on liver macrophages in patients with hepatic schistosomiasis: relationship to plasma fibronectin and circulating immunne complexes | Kh 1999 | Full text article could not be obtained |
| Sonography versus scintigraphy in chronic liver disease due to hepatitis B/schistosomiasis | Khairy 1992 | Full text article could not be obtained |
| Long-term follow-up after liver transplantation in Egyptians transplanted abroad | Khalaf 2004 | Wrong outcomes |
| Rule of schistosomiasis infection in chronic hepatitis C | Khaled 2007 | Full text article could not be obtained |
| Hepatocellular carcinoma and schistosomiasis japonica - a clinicopathologic study of 59 autopsy cases of hepatocellular carcinoma associated with chronic schistosomiasis japonica | Kojiro 1986 | No reference population given |
| Anti-HCV-positive cirrhosis associated with schistosomiasis | Koshy 1993 | Wrong outcomes |
| Chronic hepatitis B antigenaemia in bilharzial patients treated with Praziquantel | Kotkat 1990 | Insufficient information to calculate outcome effect sizes |
| The diseases that wash along with the wave of refugees challenge public health | Kreuzberg 2015 | Full text article could not be obtained |
| Is delta hepatitis infection a contributing factor for massive splenomegaly in bilharzial liver affection | Laila 1990 | Full text article could not be obtained |
| Hepatitis C virus infection in hepatic schistosomiasis | Laila 1994 | Wrong outcomes |

| | | |
|---|---|---|
| INF-gamma, IL-5 and IGE profiles in chronic schistosomiasis mansoni Egyptian patients with or without hepatitis C infection | Laila 2006 | Wrong outcomes |
| Evaluation of fibrosis sero-markers versus liver biopsy in Egyptian patients with hepatitis C and/or NASH and/or schistosomiasis | Laila 2009 | Full text article could not be obtained |
| Schistosomiasis and associated infections | Lambertucci 1998 | Review/editorial |
| Absence of relationship between Schistosoma mansoni and hepatitis B virus infection in the Qalyub Governate, Egypt | Larouze 1987 | Wrong outcomes |
| Three patients with schistosomiasis and chronic C hepatitis | Leri 1996 | Review/editorial |
| Observation on eosinophilic intranuclear inclusions in hepatocytes of patients with advanced schistosomiasis japonica | Li 1989 | Full text article could not be obtained |
| Detection of HBAg in the liver in patients with schistosomiasis complicated by liver carcinoma | Li 1990 | Wrong outcomes |
| A study and analysis of the deaths due to advanced Schistosoma japonicum infection in the Dongting Lake area of China | Li 1993 | Insufficient information to calculate outcome effect sizes |
| A clinical study on lamivudine treatment for advanced schistosomisis with chronic B hepatits | Li 2013 | No reference population given |
| Derivation and external validation of a model to predict 2-year mortality risk of patients with advanced schistosomiasis after discharge | Li 2019 | Wrong outcomes |
| Prevalence and incidence of advanced schistosomiasis and risk factors for case fatality in Hunan Province, China | Li 2021 | No reference population given |
| A research on the relationship between clinical and histological diagnosis in late stage schistosomiasis japonica | Ling 1998 | Wrong outcomes |
| Relationship between schistosomiasis and HBsAg in heavily endemic areas of islet subtype of marshland and lake regions | Liu 2006 | Full text article could not be obtained |
| Schistosome infection aggravates HCV-related liver disease and induces changes in the regulatory T-cell phenotype | Loffredo-Verde 2015 | Wrong outcomes |
| Impact of HBV and S. mansoni on portal pressure: Synergy or innocent bystanders? | Lombardi 2018 | Review/editorial |
| Long-term prognosis and risk factors of splenomegalic advanced schistosomiasis japonica | Luo 2008 | Full text article could not be obtained |
| Curative effect of anti-HBV treatment in advanced schistosomiasis patients with ascites and HIBV infection | Luo 2015 | No reference population given |
| Hepatitis B surface antigen carrier state in hepatosplenic schistosomiasis | Lyra 1976 | Wrong outcomes |
| Important risk factors related to hepatocellular carcinoma in Egypt | M 1994 | Full text article could not be obtained |
| Prevalence of antibodies to hepatitis C virus in patients with chronic liver disease | M 1994 | Wrong outcomes |
| <The> etiology of endemic hepatosplenomegaly among children | M 1995 | Full text article could not be obtained |
| Serum laminin P1 and procollagen III peptide in egyptian children with chronic hepatopathies | M 2000 | Full text article could not be obtained |
| Portal tract lymph and blood vessels changes in human chronic viral hepatitis, schistosomiasis and hepatocellular carcinoma; immunohistochemical and morphometric study | M 2005 | Full text article could not be obtained |
| Prevalence of hepatitis C infection and schistosomiasis in Egyptian patients with hepatocellular carcinoma | Mabrouk 1997 | Wrong outcomes |
| Chronic liver disease in black children in Durban, South Africa | Mackenjee 1984 | Full text article could not be obtained |
| Hepatic schistosomiasis and chronic active hepatitis | Madwar 1989 | Full text article could not be obtained |
| The relationship between uncomplicated schistosomiasis and hepatitis B infection | Madwar 1989 | Insufficient information to calculate outcome effect sizes |
| A prospective study: prediction of the first variceal haemorrhage in schistosomal and non schistosomal liver disease | Madwar 1997 | Non-eligible diagnostic method used for schistosomiasis |
| Does schistosomiasis affect cellular proliferation of hepatocytes in hepatitis C infection? | Maha 2000 | Full text article could not be obtained |
| The impact of schistosomiasis co-infection in the presentation of viral hepatitis B in migrants: An observational study in non-endemic area | Marchese 2020 | Non-endemic setting |
| Increased hepatotoxicity among HIV-infected adults co-infected with Schistosoma mansoni in Tanzania: A cross-sectional study | Marti 2017 | Wrong outcomes |
| Etiology and outcome of liver granulomatosis: A retrospective study of 21 cases | Martin-Blondel 2010 | Wrong outcomes |
| Chronic schistosomiasis japonica is an independent adverse prognostic factor for survival in hepatocellular carcinoma patients who have undergone hepatic resection: clinicopathological and prognostic analysis of 198 consecutive patients | Matsuda 2009 | Wrong outcomes |
| Control and elimination of Schistosoma mansoni infection in adult individuals on Ukerewe island, northwestern Tanzania: baseline results before implementation of intervention measures. | Mazigo 2024 | Wrong outcomes |
| Role of non invasive biomarkers in the assessment of liver condition in chronic hepatitis C Egyptian patients and if they correlate with the severity of liver affection | Mohab 2005 | No reference population given |

| | | |
|---|---|---|
| Influence of reactive oxygen species mediated immune response to Schistosomiasis and viral Hepatitis B | Mohamed 1998 | Full text article could not be obtained |
| HCV-related morbidity in a rural community of Egypt | Mohamed 2006 | Non-eligible diagnostic method used for schistosomiasis |
| Percutaneous ablation [radiofrequency and ethanol injection] versus hepatic resection in treatment of HCC in Egypt | Mohamed 2007 | Full text article could not be obtained |
| <The> role of fibroscan as a non-invasive predictor for oesophageal varices in post hcv cirrhotic egyptian patients with or without bilhariziasis | Mohamed 2018 | Insufficient information to calculate outcome effect sizes |
| Hepatitis-B virus determinants in patients with liver disease in Malawi | Molyneux 1980 | Wrong outcomes |
| Preliminary evaluation of cytokines in the hepatitis C-schistosomiasis co-infection | Morais 2006 | Insufficient information to calculate outcome effect sizes |
| Prevalence of HCV antibody and HBs antigen in schistosomal patients in relation to some risk factors | Mostapha 1995 | Full text article could not be obtained |
| Liver ultrasound findings in a low prevalence area of S. japonicum in China: comparison with history, physical examination, parasitological and serological results | Mott 1992 | Insufficient information to calculate outcome effect sizes |
| Value of alpha-smooth muscle actin and glial fibrillary acidic protein in predicting early hepatic fibrosis in chronic hepatitis C virus-infection | MoussaMohamed 2009 | Full text article could not be obtained |
| Association of HBsAg with hepatosplenic schistosomiasis II A clinico-pathological study of HBsAg and anti HBs in serum | N 1983 | Full text article could not be obtained |
| Viral hepatitis and schistosomiasis as risk factors for hepatocellular carcinoma | Nabweteme 2018 | Full text article could not be obtained |
| IL-17 Induced The Recruitment and Functional Activity of Granulocytes Isolated from Patients Coinfected with Schistosoma mansoni and Hepatitis C Virus | Nady 2017 | Wrong outcomes |
| Altered serum leptin and lipoproteins pattern in Egyptian patients with liver cirrhosis | Naglaa 2001 | Full text article could not be obtained |
| Primary liver cancer coincident with Schistosomiasis japonica. A study of 24 necropsies | Nakashima 1975 | No reference population given |
| A historical view of schistosomiasis japonica in the Chikugo river basin. What can we learn from autopsy? | Nakashima 2003 | Review/editorial |
| Abnormal liver function in different patients with Schistosoma japonicum | Ning 2014 | Wrong outcomes |
| Hepatitis-associated antigen in chronic liver diseases in Upper-Egypt | Nooman 1973 | Wrong outcomes |
| Hepatitis B virus vs. schistosomiasis and hepatocellular carcinoma in Saudi Arabia | Nouh 1990 | Wrong outcomes |
| The burden, pattern and factors that contribute to periportal fibrosis in HIV-infected patients in an S. Mansoni endemic rural Uganda | Ocama 2017 | Insufficient information to calculate outcome effect sizes |
| Clinical observations during a relatively early stage of hepatocellular carcinoma, with special reference to serum alpha-fetoprotein levels | Okuda 1975 | Wrong study design |
| Study of contact activation in endemic hepatosplenomegaly | Omran 1991 | Wrong outcomes |
| Unexplained chronic liver disease in Eastern Ethiopia: A cross-sectional study | Orlien 2016 | Wrong outcomes |
| Clinical, laboratory and liver histology of HBsAg-positive volunteer blood donors in Belo Horizonte, State of Minas Gerais, Brazil | Paz 1998 | Full text article could not be obtained |
| Hepatitis B virus infection in Schistosomiasis mansoni | Pereira 1994 | Wrong outcomes |
| Hepatitis C virus infection in schistosomiasis mansoni in Brazil | Pereira 1995 | No reference population given |
| Specific liver autoreactivity in schistosomiasis mansoni | Pereira 1997 | Wrong outcomes |
| B and C hepatitis in schistosomiasis mansoni | Pereira 2001 | Wrong outcomes |
| Cirrhosis following hepatitis C, schistosomiasis and pulmonary artery hypertension. Trial with a interferon alpha treatment | Poiraud 2000 | Full text article could not be obtained |
| Evaluation of schistosomal morbidity in subjects with high intensity infections in Qalyub, Egypt | Pope 1980 | Wrong outcomes |
| Detection of hepatitis B surface antigenaemia in patients with schistosomal hepatic fibrosis | R 1982 | Full text article could not be obtained |
| Role of HBV and HCV infections in Egyptian children and adolescents with chronic liver disease | Ragaa 1992 | Wrong outcomes |
| Impact of helicobacter pylori infection on liver pathology in egyptian patients with chronic hepatitis C | Ragheb 2011 | Full text article could not be obtained |
| Prevalence and epidemiological features of hepatocellular carcinoma in Egypt - A single center experience | RahmanEl-Zayadi 2001 | Wrong outcomes |
| Oxidative stress pattern in hepatitis C patients co-infected with schistosomiasis | Ramadan 2012 | Wrong outcomes |
| Hepatocellular carcinoma surveillance among people living with hepatitis B in Senegal (SEN-B): insights from a prospective cohort study | RamirezMena 2024 | Wrong outcomes |
| Impact of old schistosomiasis infection on the use of fibroscan for staging of fibrosis in chronic HCV patients | Ramzy 2015 | Insufficient information to calculate outcome effect sizes |
| Impact of old Schistosomiasis infection on the use of transient elastography (Fibroscan) for staging of fibrosis in chronic HCV patients. | Ramzy 2017 | Wrong outcomes |
| Immunoglobulin: a and pathogenesis of schistosomal glomerulopathy | Rashad 1995 | Wrong outcomes |
| Insulin growth factor-1 and insulin growth factor binding protein-3 in Egyptian patients with chronic hepatitis C | Raslan 2007 | Insufficient information to calculate outcome effect sizes |
| Increased tumor necrosis factor in chronic liver disease relation to severity and histopathologic pattern | Refaat 1994 | Wrong outcomes |

| | | |
|---|---|---|
| Screening for hepatocellular carcinoma among adults with HIV/HBV co-infection in Zambia: a pilot study. | Riebensahm 2022 | Wrong outcomes |
| HEPATOCELLULAR CARCINOMA SCREENING AMONG HIV/HBV-COINFECTED INDIVIDUALS IN ZAMBIA | Riebensahm 2023 | Wrong outcomes |
| Epidemiological characteristics and response to peginterferon plus ribavirin treatment of hepatitis C virus genotype 4 infection | Roulot 2007 | Wrong outcomes |
| Association of HB, Ag with hepatosplenic schistosomiasis I Relation of HBAg in liver cells, to histopathological diagnosis of liver biopsy | S 1983 | Full text article could not be obtained |
| Study on the role of schistosomiasis and hepatitis in the development of hepatocellular carcinoma | S 1993 | Full text article could not be obtained |
| Relation between hepatic lymphocyte immunophenotyping and the degree of fibrosis in schistosomal patients with and without viral B hepatitis | S 1994 | Full text article could not be obtained |
| Causes of minimal hepatic periportal fibrosis present in Egypt | S 1997 | Full text article could not be obtained |
| Kupffer cell count and plasma fibronectin level in hepatic schistosomiasis with and without virus hepatitis | S 1999 | Wrong outcomes |
| AN INITIAL INDICATION OF PREDISPOSING RISK OF SCHISTOSOMA MANSONI INFECTION FOR HEPATOCELLULAR CARCINOMA | SabryAe 2015 | No reference population given |
| Response to hepatitis b vaccine in chronic hepatitis C Egyptian patients | Said 2015 | Full text article could not be obtained |
| Clearance of hepatitis C virus infection did not ameliorate the response to hepatitis B vaccine | Said 2022 | Full text article could not be obtained |
| Study of the cytokine transforming growth factor beta-1 in patients with chronic virus hepatitis | Samir 2003 | Full text article could not be obtained |
| C-MYC oncogen overexpression in Egyptian patients with hepatocellular Carcinoma: Association with viral and/or Schistosoma mansoni infection | Samir 2011 | Full text article could not be obtained |
| Studies on relationship between schistosomiasis and viral hepatitis infection | SamirAbdelRazik 2006 | Full text article could not be obtained |
| Neglected tropical diseases as a cause of chronic liver disease: The case of Schistosomiasis and Hepatitis C Co-infections in Egypt | Sanghvi 2013 | Review/editorial |
| <A> modified warren shunt by using 8-mm interposition Gore-Tex graft | Sarwat 1991 | Full text article could not be obtained |
| Clinical significance of urinary neopterin in chronic liver disease | Shahira 1998 | Wrong outcomes |
| Low dose splenic irradiation for the treatment of hypersplenism in compensated and decompensated liver cirrhosis | Sheir 2005 | Wrong outcomes |
| Hepatocellular carcinoma: A clinicopathological analysis of 118 cases from Riyadh Central Hospital | Sherbini 1992 | Wrong outcomes |
| Immunological studies on liver in Egyptian infants and children | Sherif 1986 | Wrong outcomes |
| Comparison between transient elastography (Fibroscan) and liver biopsy for diagnosis of hepatic fibrosis in chronic hepatitis C genotype 4 | Shiha 2012 | Full text article could not be obtained |
| Schistosoma mansoni infection and the occurrence, characteristics, and survival of patients with hepatocellular carcinoma: an observational study over a decade | Shousha 2022 | Insufficient information to calculate outcome effect sizes |
| Analysis of coagulation related parameters between patients with advanced schistosomiasis cirrhosis and hepatitis B cirrhosis | Shun 2016 | Wrong outcomes |
| Hepatitis-B virus infection among Yemeni patients with hepatocellular carcinoma | Silm 2004 | Non-eligible diagnostic method used for schistosomiasis |
| Assessing Thrombocytopenia and Chronic Liver Disease in Southeast Asia: A Multicentric Cross-Sectional Study. | Sohail 2023 | Wrong outcomes |
| Study of some associated factors with hepatocellular carcinoma in Egypt | Soliman 2011 | Full text article could not be obtained |
| Immunohistochemical study of the relationship between advanced hepatosplenic schistosomiasis japonica and viral hepatitis | Song 1985 | Wrong outcomes |
| AETIOLOGY AND OUTCOMES OF CIRRHOSIS AND HEPATOCELLULAR CARCINOMA IN BLANTYRE, MALAWI | Stockdale 2023 | Full text article could not be obtained |
| Clinical characteristics and response to therapy in Egyptian children heavily infected with Schistosoma mansoni | Strickland 1982 | Wrong outcomes |
| Role of hepatitis C infection in chronic liver disease in Egypt | Strickland 2002 | Wrong outcomes |
| Hepatocellular carcinoma: a clinicopathological analysis of 118 cases from Riyadh Central Hospital | Suphia 1992 | Full text article could not be obtained |
| Clinicoepidemiological characteristics and response to treatment in patients with hepatocellular carcinoma in Egypt | Taha 2012 | Full text article could not be obtained |
| Intercellular adhesion molecule-1 in sera of patients with chronic viral hepatitis | TahaniAhmad 2001 | Full text article could not be obtained |
| Serological markers of hepatitis B and C in residents of a schistosomiasis endemic area | Tavares-Neto 1998 | Full text article could not be obtained |
| Clinical regression of the hepatosplenic form of schistosomiasis mansoni and serological markers for hepatitis B virus | Tavares-Neto 2005 | No reference population given |
| Liver surgery for hepatocellular carcinoma (HCC) in nonalcoholic steatohepatitis (NASH): A single-center experience | Teixeira 2017 | Full text article could not be obtained |
| The relation of schistosomiasis complicated by hepatitis B and primary cancer of the liver | Tu 1989 | Full text article could not be obtained |

| | | |
|---|---|---|
| Antibody to hepatitis C virus in patients with chronic schistosomiasis | Uemura 1992 | Full text article could not be obtained |
| Transient elastography evaluation of hepatic and spleen stiffness in patients with hepatosplenic schistosomiasis | Veiga 2015 | Wrong outcomes |
| Renal health after long-term exposure to tenofovir disoproxil fumarate (TDF) in HIV/HBV positive adults in Ghana | Villa 2018 | Insufficient information to calculate outcome effect sizes |
| Validity of platelet count/spleen diameter ratio as a predictor of esophageal varices in patients with liver cirrhosis | Wafaa 2008 | Wrong outcomes |
| Cell mediated immune response in chronic liver diseases: schistosomal, viral and neoplastic | Wahib 1998 | Full text article could not be obtained |
| Carcinoembryonic antigen in schistosomiasis | Watsky 1982 | Review/editorial |
| Hepatic parenchymal dysfunction in Schistosoma japonicum infection | Watt 1991 | Wrong outcomes |
| The etiology of liver damage imparts cytokines transforming growth factor beta1 or interleukin-13 as driving forces in fibrogenesis | Weng 2009 | Wrong outcomes |
| The diagnosis and treatment of small liver cancer | Wenhe 1999 | Full text article could not be obtained |
| Morbidity due to schistosiomiasis japonica in the People's Republic of China | Wiest 1992 | Wrong outcomes |
| Pathologic study on 53 cases of schistosomiasis japonica with hepatitis B | Wu 1983 | Wrong outcomes |
| Analysis of HBV infection in patients with chronic schistosomiasis and advanced schistosomiasis | Xu 2005 | Full text article could not be obtained |
| Clinical investigation on patients with advanced schistosomiasis and HBV infection | Yao 2005 | Wrong outcomes |
| A case-control study on risk factors for advanced schistosomiasis japonica | Yuan 2002 | Full text article could not be obtained |
| Predictive value of the red blood cell distribution width-to-platelet ratio for hepatic fibrosis | Yuyun 2019 | Wrong outcomes |
| Prevalence of HBs-Ag in schistosomiasis: B-frequency in various stages of schistosomiasis | Zakaria 1979 | Full text article could not be obtained |
| The role of schistosomiasis and type B hepatitis in the pathogenesis of endemic Egyptian hepatosplenomegaly | Zakaria 1988 | Insufficient information to calculate outcome effect sizes |
| Evaluation of advanced liver disease with ascites by sonography, histopathology and serology | Zakaria 1989 | Insufficient information to calculate outcome effect sizes |
| T helper and T suppressor cells in schistosomal and non-schistosomal chronic liver disease | Zakaria 1993 | Wrong outcomes |
| Value of a-smooth muscle actin and glial fibrillary acidic protein in predicting early hepatic fibrosis in chronic hepatitis C virus infection | Zakaria 2010 | Insufficient information to calculate outcome effect sizes |
| Morbidity of schistosomiasis mansoni in rural Alexandria, Egypt | Zaki 2003 | Wrong outcomes |
| Study of the association of hepatitis B surface antigen and HLA with hepatocellular carcinoma | Zaki 2010 | Full text article could not be obtained |
| Histocompatibility antigens in relation to hepatic schistosomiasis | Zaki 2010 | Full text article could not be obtained |
| Study of the cellular immunity in patients with hepatic schistosomiasis with and without hepatitis C viral infection | Zaki 2011 | Full text article could not be obtained |
| Cytokine profile in Egyptian hepatitis C virus genotype-4 in relation to liver disease progression | Zekri 2005 | Wrong outcomes |
| Detection and significance of HBV-DNA and/or HCV-RNA in sera and livers of advanced schistosomiasis cases | Zhang 1997 | Wrong outcomes |
| Observation of T lymphocyte subsets in the liver of patients with advanced schistosomiasis and advanced schistosomiasis accompanied with hepatitis B | Zhang 2000 | No reference population given |
| Effects of HBV infection on hepatic fibrosis and level of Th1/Th2 cytokines in the patients with Schistosomiasis japonica | Zhong-WeiJIA 2003 | Full text article could not be obtained |
| Etiological and clinicopathologic characteristics of intrahepatic cholangiocarcinoma in young patients | Zhou 2010 | Wrong outcomes |
| Hepatitis B virus-associated intrahepatic cholangiocarcinoma and hepatocellular carcinoma may hold common disease process for carcinogenesis | Zhou 2010 | Wrong outcomes |

### Table S10: Variables extracted from studies and their definitions

| Variable | Definition |
|---|---|
| **ID** | covidence ID/study ID |
| **title** | Title of paper |
| **author** | Last name of first author |
| **publication_year** | Year of publication |
| **citation** | Citation (Vancouver format) |
| **study_year** | Year of study (year of outcome measurement) |
| **studydur_months** | Total duration of study in months |
| **country** | Study country |
| **study_area** | Study area where the exact administrative unit is recorded as provided by the study authors to the lowest administrative level (village name, district name, etc.) |
| **locality** | Rural - e.g. villages, remote areas away from built-up regions<br>Peri-urban - outskirts of a city/town, on the rural-urban fringe<br>Urban - e.g. cities/towns, densely-population and built-up |
| **study_setting_locality** | Location where study was conducted: community, school, health clinic, community + school, community + health clinic, school + health clinic, all three |
| **sampling_strategy** | Method by which study participants were identified and enrolled (refers to sampling method to analyse association between water contact and infection, this may be subgroup of larger study population). Options: convenience, random, systematic, snowball, stratified convenience, stratified random, stratified systematic, stratified snowball, not reported |
| **inclusion** | Study inclusion criteria for participants |
| **exclusion** | Study exclusion criteria for participants |
| **aim** | Main study aim (as defined by authors) |
| **design** | Type of study (cross-sectional study, before-after study, randomised controlled trial, cohort study) |
| **sample_size** | Total number of participants recruited (participants with both infection and liver fibrosis information) |
| **age_cat** | Summary of included age groups. Options: PSAC, SAC, Older children & Adults (multiple-select). Definitions used as provided by study authors. If none were provided then age groups were assigned (PSAC (0-4 years), SAC (5-14 years), Older children and Adults (15+ years) |
| **age_lower** | Lower limit of participant age range |
| **age_upper** | Upper limit of participant age range |
| **gender** | Summary of participant gender. Options: male, female, both |
| **schisto_species** | Schistosoma species. Options: *S. japonicum*, *S. mansoni*, *S. mekongi* |
| **schisto_diagnostic** | Diagnostic test category. Options: Microscopy, antigen, antibody. |
| **schisto_diagnostic_exact** | Specific diagnostic test used. Options: Kato-Katz, POC-CCA, PCR, other as defined by study authors |
| **schisto_infection_status_def** | Definition of infection status as provided by study authors |
| **schisto_infection_intensity_def** | Definition of infection intensity as provided by study authors |
| **schisto_infected_total** | Number of participants that are infected |
| **hep_species** | B or C |
| **hepB_diagnostic** | In case of hep B positivity - exact diagnostic method used: antigen/antibody/viral load |
| **hepC_diagnostic** | In case of hep C positivity - exact diagnostic method used: antigen/antibody/viral load |

| | |
|---|---|
| **hep_chronic** | >6 months HBC RNA presence, >6 months HBsAg positive |
| **hep_infection_status_def** | Definition of infection status as provided by study authors |
| **hep_infection_intensity_def** | Definition of infection intensity as provided by study authors |
| **hep_infected_total** | Number of participants that are infected with either Hep B or Hep C |
| **hep_coinfected_total** | Number of participants coinfected with hep B + hep C (regardless of Schistosomiasis infection status) |
| **coinfected_total** | Number of participants with coinfections (hep B or C + one of *S. mansoni, S. japonicum, S. mekongi*) |
| **path_total** | Number of participants with any of the four liver pathologies of interest |
| **pathi** | Number of participants with coinfection (hep B or C + one of *S. mansoni, S. japonicum, S. mekongi*) and any of the four liver pathologies of interest |
| **nopathi** | Number of participants with coinfection (hep B or C + one of *S. mansoni, S. japonicum, S. mekongi*) and no liver pathology |
| **pathn** | Number of participants singularly or non-infected with any of the four liver pathologies of interest |
| **nopathn** | Number of participants singularly or non-infected with no liver pathology |
| **path_OR** | Odds ratio or risk given (or converted) for any pathology with co-infection |
| **pa95ci** | 95% confidence interval for OR given for any pathology present in event of co-infection |
| **HM_total** | Total (%) of study population who have hepatomegaly as a liver outcome |
| **HM_coinfect** | Individual percentage or number of study population with coinfection + hepatomegaly |
| **noHM_coinfect** | Individual percentage or number of study population with coinfection + no hepatomegaly |
| **HM_noninfect** | Individual percentage or number of study population with no infection OR singular infection + hepatomegaly |
| **noHM_noninfect** | Individual percentage or number of study population with no infection OR singular infection + no hepatomegaly |
| **HM_OR** | Odds ratio or risk given (or converted) for hepatomegaly with co-infection |
| **HM_95CI** | 95% confidence interval for OR given for hepatomegaly present in event of co-infection |
| **HM_diag** | Descriptive (author-defined) tool/method used to diagnose hepatomegaly (e.g. clinical examination, ultrasonography) |
| **HM_authordef** | Author definition of hepatomegaly |
| **fibrosis_total** | Total (%) of study population who have liver fibrosis as a liver outcome |
| **fibrosis_coinfect** | Individual percentage or number of study population with coinfection + fibrosis |
| **nofibrosis_coinfect** | Individual percentage or number of study population with coinfection + no fibrosis |
| **fibrosis_noninfect** | Individual percentage or number of study population with no infection OR singular infection + fibrosis |

| | |
|---|---|
| **nofibrosis_noninfect** | Individual percentage or number of study population with no infection OR singular infection + no fibrosis |
| **fibrosis_OR** | Odds ratio or risk given (or converted) for fibrosis with co-infection |
| **fibrosis_95CI** | 95% confidence interval for OR given for fibrosis present in the event of co-infection |
| **fibrosis_diag** | Descriptive (author-defined) tool/method used to diagnose fibrosis (e.g. serum markers, liver biopsy, imaging) |
| **fibrosis_authordef** | Author definition of fibrosis |
| **cirrhosis_total** | Total (%) of study population who have liver cirrhosis as a liver outcome |
| **cirrhosis_coinfect** | Individual percentage or number of study population with coinfection + cirrhosis |
| **nocirrhosis_coinfect** | Individual percentage or number of study population with coinfection + no cirrhosis |
| **cirrhosis_noninfect** | Individual percentage or number of study population with no infection OR singular infection + cirrhosis |
| **nocirrhosis_noninfect** | Individual percentage or number of study population with no infection OR singular infection + no cirrhosis |
| **cirrhosis_OR** | Odds ratio or risk given (or converted) for cirrhosis with co-infection |
| **cirrhosis_95CI** | 95% confidence interval for OR given for cirrhosis present in the event of co-infection |
| **cirrhosis_diag** | Descriptive (author-defined) tool/method used to diagnose cirrhosis (e.g. liver biopsy, imaging, liver function tests) |
| **cirrhosis_authordef** | Author definition of cirrhosis |
| **HCC_total** | Total (%) of study population who have hepatocellular carcinoma as a liver outcome |
| **HCC_coinfect** | Individual percentage or number of study population with coinfection + HCC |
| **noHCC_coinfect** | Individual percentage or number of study population with coinfection + no HCC |
| **HCC_noninfect** | Individual percentage or number of study population with no infection OR singular infection + HCC |
| **nocirrhosis_noninfect** | Individual percentage or number of study population with no infection OR singular infection + no HCC |
| **HCC_OR** | Odds ratio or risk given (or converted) for HCC with co-infection |
| **HCC_95CI** | 95% confidence interval for OR given for HCC present in the event of co-infection |
| **HCC_diag** | Descriptive (author-defined) tool/method used to diagnose HCC (e.g. imaging, biopsy) |
| **HCC_authordef** | Author definition of heptocellular carcinoma |
| **mortality_total** | Total number of participants who died during study or follow up (%) |
| **HIV** | total participants (%) with current HIV infection |
| **TB** | total participants (%) with current TB infection |
| **malaria** | total participants (%) with current malaria infection |
| **prev_PZQ** | total participants (%) who have previously received any schistosomal treatment (perenteral or praziquantel) |

| | |
|---|---|
| **prev_hepB_tx** | total participants (%) who have previously received any hepatitis B treatment incl hep B vaccine |
| **prev_hepC_tx** | total participants (%) who have previously received any hepatitis C treatment |
| **hepatosplen_otherpath** | list any other reported hepatic or splenic pathologies given by study authors in the context of co-infection. Where any others are available, to create a new column for each with % of participants in study population. |

A summarised version of the data extraction table is included as Appendix 2.

**Table S11: Risk of bias assessment tool adapted from National Institutes of Health Tool from the National Heart, Lung, and Blood Institute for observational cohort and cross-sectional studies.**

| Category | Questions | Guidance on when to select 'yes' | Guidance on when to select 'no' |
|---|---|---|---|
| **Aim** | **Q1) Was the research question or objective in this paper clearly stated?** | Authors clearly describe the goal of their research, e.g. the aim was to estimate the association between *Schistosoma mansoni/S. japonicum/S. mekongi* and hepatitis B or C co-infection status on liver pathology. | No aim, unspecific aim, aim does not match the analyses completed or aim does not match the outcomes reported. |
| **Representativeness** | **Q2) Was the study population clearly specified and defined?** | Authors describe the group of people from which the study participants were selected or recruited, using demographics, location, and time period. | Description lacking specifics on study population demographics, location, and time period. |
| | **Q3) Was the method for selecting the sample clearly described?** | Sampling method clearly described, e.g. 10 school-age children (5-14 years old) per class in each of the five schools were sampled using stratified random sampling. | Lacking details to understand how sampling was done, e.g. 50 adults per village were selected for inclusion in the study. |
| | **Q4) Was the participation rate of eligible persons at least 50%?** | Provides statistics on response rate, e.g. 90% of responders selected from village registries agreed to participate in the study. | Low response rate/no information given. |

| | | | |
|---|---|---|---|
| | **Q5) Is the sample representative of the population from which it is drawn (including similar timeframe)?**<br><br>**Q6) Were inclusion and exclusion criteria for being in the study pre-specified and applied uniformly to all participants? (For case-control studies: Were the cases clearly defined and differentiated from controls?)** | Clear inclusion and exclusion criteria. Selected participants representative of the population they are sampled from (and in cohort or case-control studies, both groups selected from same underlying population), e.g. random sample from community.<br><br>(For case-control studies: cases and controls are clearly defined, cases and controls are drawn from the same population and there are no significant differences between the cases and controls). | Sample not representative of population, e.g. children sampled in schools may not be representative of school-age children, or participants selected based on liver fibrosis may not be representative of village population (e.g. higher social status households, all households with home latrines, etc.). Excluded participants without providing a reason.<br><br>(For case-control studies: no clear definitions of cases and/or controls, cases and controls are not matched or stratified, despite being specified in the design). |
| | **Q7) Was a sample size justification, power description, or variance and effect estimates provided?** | Authors present their reasons for selecting or recruiting the number of people included or analysed, e.g. state that they performed power calculations before selecting sample. | No justification on size of selected sample provided. |
| **Exposure** | **Q8) Were the exposures of interest measured prior to the outcome being measured?**<br><br>**Q9) Was the timeframe sufficient so that one could reasonably expect to see an association between exposure and outcome if it existed?** | Information relating to liver pathology was obtained post infection status being obtained, i.e. the investigator can confirm that the exposure (coinfection) occurred prior to the development of hepatic pathology. | The investigator cannot confirm that the infection occurred prior to the development of hepatic pathology, e.g. authors measured infection status after the detection of liver fibrosis by ultrasound. |

| | | | |
|---|---|---|---|
| | **Q10) Did the study define the exposures?** | The study describes infection status of *S. mansoni, S. japonicum or S. mekongi* eggs seen on stool microscopy using the Kato Katz technique, or POC-CCA positive with a form of quality control measure, e.g. ensuring a senior laboratory technician re-analyses 10% of stool sample using the Kato-Katz microscopy technique. For the use of a commercial diagnostic for antigen-based and antibody-based tests, analyses should be done in a reference laboratory. Infection status for hepatitis B and C should clearly be defined as antigenic or antibody positivity. | Study does not explicitly define infection status. |
| | **Q11) Were the assessors of exposure blinded to the case or control status of participants?** | Individuals performing diagnostic modalities (imaging, phlebotomy, clinical examination) are different to those interpreting findings (e.g. radiologists, lab technicians). | Individuals performing diagnostic modalities (imaging, phlebotomy, clinical examination) are the same as those analysis results. |

| | | | |
|---|---|---|---|
| **Outcomes** | **Q12) Were the outcomes clearly defined, valid, reliable, and implemented consistently across all study participants?** | Study clearly states diagnosis of recognised hepatic pathology using eligible diagnostic modalities:<br><br>Height-standardised hepatomegaly diagnosed by abdominal palpation or imaging; liver fibrosis diagnosed by recognised and approved serum biomarkers, biopsy, or imaging; liver cirrhosis diagnosed by recognised and approved serum biomarkers, biopsy, or imaging; and hepatocellular carcinoma diagnosed by serum biomarkers (only in conjunction with liver imaging and/or biopsy), biopsy, or imaging.<br><br>Use of eligible scoring systems for grading liver fibrosis (e.g. APRI, Bonacini index, Fib-4, ELF index, Fibro index).<br><br>Authors explicitly state which scoring system for grading liver fibrosis on biopsy was used (e.g. Ishak score, Metavir). This system can either be a previously clinically accepted system, or a newly developed and validated system. | Study does not state use of eligible diagnostic method for diagnosis of recognised hepatic pathology.<br><br>Study does not state eligible grading system for assessing grade of fibrosis or cirrhosis. |
| | **Q13) Were exposures and outcomes not measured by the same person, i.e. were the outcome assessors blinded to the exposure status of participants?** | Hepatitis B/C and schistosomiasis (co-) infection status not known by individual assessing liver (e.g. different person performing blood sample analysis and ultrasound sonography/interpreting findings). | Hepatitis B/C and schistosomiasis (co-) infection status known by individual assessing liver pathology (e.g. same technician analysis blood samples and performing ultrasound sonography/interpreting findings). |
| | **Q14) Was loss to follow-up after baseline 20% or less? Is there missing data?** | Follow-up in studies with multiple rounds of data collection not less than 80%, e.g. attrition at follow-up was 5%. | Loss to follow-up was 20% or more. Or if there were 200 participants in the study but only 100 in the model, missingness is >20% and therefore unacceptable. No information on loss to follow-up given. |

| | | | |
|---|---|---|---|
| **Covariates** | **Q15) Were the covariates clearly defined, valid, reliable, and implemented consistently across all study participants?** | For example, age categories 5-14 years, 15-18 years, 18+ years given. | Covariates lacking information on definition. |
| **Analysis** | **Q16) Were adjusted (inclusive of at least age and gender) and unadjusted effect measures and their 95% CIs reported?** | Studies report both types of models and report on the covariates adjusted for. | Studies report only adjusted or unadjusted ORs/RRs, fail to provide 95% CIs or descriptions of variables adjusted for. |

### Table S12: Full risk of bias assessment of included studies

| | Q1 | Q2 | Q3 | Q4 | Q5 | Q6 | Q7 | Q8 | Q9 | Q10 | Q11 | Q12 | Q13 | Q14 | Q15 | Q16 | S | I |
|---|---|---|---|---|---|---|---|---|---|---|---|---|---|---|---|---|---|---|
| Abbas, 2009 | 1; Authors clearly describe the goal of their research | 1; Clear description of the study population using demographics, location and time period. | 0; Lacking details to understand how participant sampling was performed | 0; No information given | 0; Unable to determine | 1; Clear inclusion and exclusion criteria given. | 0; No justification on size of selected sample provided. | 1; Exposures of interest were measured prior to the outcome. | 0; No information given. | 1; Clear explanation of exposure of interest and how this was determined. | 0; No information given. | 1; Study clearly states diganosis of recognised hepatic pathology using eligible diagnostic modality. | 0; No information given. | 0; No information given. | 0; No information on covariates given. | 1; Yes, model provided. | 7 | M |
| Abdel-Aziz, 2012 | 1; Authors clearly describe the goal of their research | 1; Clear description of the study population using demographics, location and time period. | 1; Study describes the method of selecting the participant sample. | 0; No information given | 1; Selected participants representative of the population they are sampled from | 1; Clear inclusion and exclusion criteria given. | 0; No justification on size of selected sample provided. | 1; Exposures of interest were measured prior to the outcome. | 0; No information given. | 1; Clear explanation of exposure of interest and how this was determined. | 0; No information given. | 1; Study clearly states diganosis of recognised hepatic pathology using eligible diagnostic modality. | 0; No information given. | 0; No information given. | 0; No information on covariates given. | 0; No adjusted/unadjusted models reported. | 8 | M |

| | | | | | | | | | | | | | | | | | | |
|---|---|---|---|---|---|---|---|---|---|---|---|---|---|---|---|---|---|---|
| AbdEl-Moneim, 2008 | 1; Authors clearly describe the goal of their research | 1; Clear description of the study population using demographics, location and time period. | 0; Lacking details to understand how participant sampling was performed | 0; No information given | 0; Unable to determine | 1; Clear inclusion and exclusion criteria given. | 0; No justification on size of selected sample provided. | 1; Exposures of interest were measured prior to the outcome. | 0; No information given. | 1; Clear explanation of exposure of interest and how this was determined. | 0; No information given. | 1; Study clearly states diganosis of recognised hepatic pathology using eligible diagnostic modality. | 0; No information given. | 0; No information given. | 0; No information on covariates given. | 0; No adjusted/unadjusted models reported. | 6 | M |
| Ahmed, 2015 | 1; Authors clearly describe the goal of their research | 1; Clear description of the study population using demographics, location and time period. | 0; Lacking details to understand how participant sampling was performed | 0; No information given | 0; Unable to determine | 1; Clear inclusion and exclusion criteria given. | 0; No justification on size of selected sample provided. | 1; Exposures of interest were measured prior to the outcome. | 1; Timeframe sufficient so an association could be seen if it existed. | 1; Clear explanation of exposure of interest and how this was determined. | 0; No information given. | 1; Study clearly states diganosis of recognised hepatic pathology using eligible diagnostic modality. | 0; No information given. | 0; No information given. | 0; No information on covariates given. | 0; No adjusted/unadjusted models reported. | 7 | M |
| Allam, 2014 | 1; Authors clearly describe the goal of their research | 1; Clear description of the study population using demographics, location and time period. | 1; Study describes the method of selecting the participant sample. | 0; No information given | 1; Selected participants representative of the population they are sampled from | 0; No clear information on inclusion and exclusion criteria given. | 0; No justification on size of selected sample provided. | 1; Exposures of interest were measured prior to the outcome. | 0; No information given. | 1; Clear explanation of exposure of interest and how this was determined. | 0; No information given. | 0; Study clearly states diagnostic method used for liver pathology, but no clear definitions or scoring systems for liver pathology were given. | 0; No information given. | 0; No information given. | 0; No information on covariates given. | 0; No adjusted/unadjusted models reported. | 6 | Mo |

| | | | | | | | | | | | | | | | | | |
|---|---|---|---|---|---|---|---|---|---|---|---|---|---|---|---|---|---|
| Allam, 2024 | 1; Authors clearly describe the goal of their research | 1; Clear description of the study population using demographics, location and time period. | 0; Lacking details to understand how participant sampling was performed | 0; No information given | 0; Unable to determine | 1; Clear inclusion and exclusion criteria given. | 0; No justification on size of selected sample provided. | 0; Exposures of interest were measured after the outcome was established. | 0; No information given. | 1; Clear explanation of exposure of interest and how this was determined. | 0; No information given. | 0; Study does not provide clear explanations on the diagnostic methods used for the outcome of interest. | 0; No information given. | 0; No information given. | 0; No information on covariates given. | 0; No adjusted/unadjusted models reported. | 5 | H |
| Amal, 2001 | 1; Authors clearly describe the goal of their research | 1; Clear description of the study population using demographics, location and time period. | 1; Study describes the method of selecting the participant sample. | 0; No information given | 1; Selected participants representative of the population they are sampled from | 0; No clear information on inclusion and exclusion criteria given. | 0; No justification on size of selected sample provided. | 1; Exposures of interest were measured prior to the outcome. | 0; No information given. | 1; Clear explanation of exposure of interest and how this was determined. | 0; No information given. | 1; Study clearly states diganosis of recognised hepatic pathology using eligible diagnostic modality. | 0; No information given. | 0; No information given. | 0; No information on covariates given. | 0; No adjusted/unadjusted models reported. | 7 | M |
| Badra, 2007 | 1; Authors clearly describe the goal of their research | 1; Clear description of the study population using demographics, location and time period. | 0; Lacking details to understand how participant sampling was performed | 0; No information given | 0; Unable to determine | 1; Clear inclusion and exclusion criteria given. | 0; No justification on size of selected sample provided. | 1; Exposures of interest were measured prior to the outcome. | 0; No information given. | 1; Clear explanation of exposure of interest and how this was determined. | 0; No information given. | 1; Study clearly states diganosis of recognised hepatic pathology using eligible diagnostic modality. | 0; No information given. | 0; No information given. | 0; No information on covariates given. | 0; No adjusted/unadjusted models reported. | 6 | M |

| | | | | | | | | | | | | | | | | | |
|---|---|---|---|---|---|---|---|---|---|---|---|---|---|---|---|---|---|
| Chen, 2009 | 1; Authors clearly describe the goal of their research | 1; Clear description of the study population using demographics, location and time period. | 0; Lacking details to understand how participant sampling was performed | 0; No information given | 0; Unable to determine | 0; No clear information on inclusion and exclusion criteria given. | 0; No justification on size of selected sample provided. | 1; Exposures of interest were measured prior to the outcome. | 0; No information given. | 1; Clear explanation of exposure of interest and how this was determined. | 0; No information given. | 1; Study clearly states diganosis of recognised hepatic pathology using eligible diagnostic modality. | 0; No information given. | 0; No information given. | 0; No information on covariates given. | 0; No adjusted/unadjusted models reported. | 5 | H |
| el-Masry, 2006 | 1; Authors clearly describe the goal of their research | 0; Description of the study population lacking specifics of demographics and time period | 0; Lacking details to understand how participant sampling was performed | 0; No information given | 0; Unable to determine | 1; Clear inclusion and exclusion criteria given. | 0; No justification on size of selected sample provided. | 1; Exposures of interest were measured prior to the outcome. | 0; No information given. | 1; Clear explanation of exposure of interest and how this was determined. | 0; No information given. | 1; Study clearly states diganosis of recognised hepatic pathology using eligible diagnostic modality. | 0; No information given. | 0; No information given. | 0; No information on covariates given. | 0; No adjusted/unadjusted models reported. | 5 | H |
| El-Shazly, 1994 | 1; Authors clearly describe the goal of their research | 0; Description of the study population lacking specifics of demographics and time period | 0; Lacking details to understand how participant sampling was performed | 0; No information given | 0; Unable to determine | 1; Clear inclusion and exclusion criteria given. | 0; No justification on size of selected sample provided. | 1; Exposures of interest were measured prior to the outcome. | 0; No information given. | 0; Study lacks specifics on how exposure was determined. | 0; No information given. | 1; Study clearly states diganosis of recognised hepatic pathology using eligible diagnostic modality. | 0; No information given. | 0; No information given. | 0; No information on covariates given. | 0; No adjusted/unadjusted models reported. | 4 | H |

| | | | | | | | | | | | | | | | | | |
|---|---|---|---|---|---|---|---|---|---|---|---|---|---|---|---|---|---|
| Emam, 2006 | 1; Authors clearly describe the goal of their research | 0; Description of the study population lacking specifics of demographics and time period | 0; Lacking details to understand how participant sampling was performed | 0; No information given | 0; Unable to determine | 1; Clear inclusion and exclusion criteria given. | 0; No justification on size of selected sample provided. | 1; Exposures of interest were measured prior to the outcome. | 0; No information given. | 1; Clear explanation of exposure of interest and how this was determined. | 0; No information given. | 1; Study clearly states diganosis of recognised hepatic pathology using eligible diagnostic modality. | 0; No information given. | 0; No information given. | 0; No information on covariates given. | 0; No adjusted/unadjusted models reported. | 5 | H |
| Esmat, 2013 | 1; Authors clearly describe the goal of their research | 0; Description of the study population lacking specifics of demographics and time period | 0; Lacking details to understand how participant sampling was performed | 0; No information given | 0; Unable to determine | 1; Clear inclusion and exclusion criteria given. | 0; No justification on size of selected sample provided. | 1; Exposures of interest were measured prior to the outcome. | 0; No information given. | 1; Clear explanation of exposure of interest and how this was determined. | 0; No information given. | 1; Study clearly states diganosis of recognised hepatic pathology using eligible diagnostic modality. | 0; No information given. | 0; No information given. | 0; No information on covariates given. | 0; No adjusted/unadjusted models reported. | 5 | H |
| Gobert, 2015 | 1; Authors clearly describe the goal of their research | 1; Clear description of the study population using demographics, location and time period. | 1; Study describes the method of selecting the participant sample. | 0; No information given | 1; Selected participants representative of the population they are sampled from | 1; Clear inclusion and exclusion criteria given. | 0; No justification on size of selected sample provided. | 1; Exposures of interest were measured prior to the outcome. | 0; No information given. | 1; Clear explanation of exposure of interest and how this was determined. | 0; No information given. | 1; Study clearly states diganosis of recognised hepatic pathology using eligible diagnostic modality. | 0; No information given. | 0; No information given. | 0; No information on covariates given. | 0; No adjusted/unadjusted models reported. | 8 | M |

| | | | | | | | | | | | | | | | | | | |
|---|---|---|---|---|---|---|---|---|---|---|---|---|---|---|---|---|---|---|
| Gunda, 2020 | 1; Authors clearly describe the goal of their research | 1; Clear description of the study population using demographics, location and time period. | 1; Study describes the method of selecting the participant sample. | 0; No information given | 1; Selected participants representative of the population they are sampled from | 0; No clear information on inclusion and exclusion criteria given. | 1; Justification on size of selected sample provided. | 1; Exposures of interest were measured prior to the outcome. | 0; No information given. | 1; Clear explanation of exposure of interest and how this was determined. | 0; No information given. | 1; Study clearly states diganosis of recognised hepatic pathology using eligible diagnostic modality. | 0; No information given. | 0; No information given. | 0; No information on covariates given. | 1; Yes, model provided. | 9 | M |
| Hassan, 2003 | 1; Authors clearly describe the goal of their research | 1; Clear description of the study population using demographics, location and time period. | 0; Lacking details to understand how participant sampling was performed | 0; No information given | 0; Unable to determine | 0; No clear information on inclusion and exclusion criteria given. | 0; No justification on size of selected sample provided. | 1; Exposures of interest were measured prior to the outcome. | 0; No information given. | 1; Clear explanation of exposure of interest and how this was determined. | 0; No information given. | 1; Study clearly states diganosis of recognised hepatic pathology using eligible diagnostic modality. | 0; No information given. | 0; No information given. | 0; No information on covariates given. | 0; No adjusted/unadjusted models reported. | 5 | H |
| Helal, 1998 | 1; Authors clearly describe the goal of their research | 1; Clear description of the study population using demographics, location and time period. | 0; Lacking details to understand how participant sampling was performed | 0; No information given | 0; Unable to determine | 1; Clear inclusion and exclusion criteria given. | 0; No justification on size of selected sample provided. | 1; Exposures of interest were measured prior to the outcome. | 0; No information given. | 1; Clear explanation of exposure of interest and how this was determined. | 0; No information given. | 1; Study clearly states diganosis of recognised hepatic pathology using eligible diagnostic modality. | 0; No information given. | 0; No information given. | 0; No information on covariates given. | 0; No adjusted/unadjusted models reported. | 6 | M |

| | | | | | | | | | | | | | | | | | | |
|---|---|---|---|---|---|---|---|---|---|---|---|---|---|---|---|---|---|---|
| Iida, 1999 | 1; Authors clearly describe the goal of their research | 1; Clear description of the study population using demographics, location and time period. | 0; Lacking details to understand how participant sampling was performed | 0; No information given | 0; Unable to determine | 1; Clear inclusion and exclusion criteria given. | 0; No justification on size of selected sample provided. | 1; Exposures of interest were measured prior to the outcome. | 0; No information given. | 1; Clear explanation of exposure of interest and how this was determined. | 0; No information given. | 0; Study does not provide clear explanations on the diagnostic methods used for the outcome of interest. | 0; No information given. | 0; No information given. | 0; No information on covariates given. | 0; No adjusted/unadjusted models reported. | 5 | H |
| Kamal, 2000 | 1; Authors clearly describe the goal of their research | 1; Clear description of the study population using demographics, location and time period. | 1; Study describes the method of selecting the participant sample. | 0; No information given | 1; Selected participants representative of the population they are sampled from | 1; Clear inclusion and exclusion criteria given. | 0; No justification on size of selected sample provided. | 1; Exposures of interest were measured prior to the outcome. | 0; No information given. | 1; Clear explanation of exposure of interest and how this was determined. | 0; No information given. | 1; Study clearly states diganosis of recognised hepatic pathology using eligible diagnostic modality. | 0; No information given. | 0; No information given. | 0; No information on covariates given. | 0; No adjusted/unadjusted models reported. | 8 | M |
| Kamal, 2001 | 1; Authors clearly describe the goal of their research | 1; Clear description of the study population using demographics, location and time period. | 1; Study describes the method of selecting the participant sample. | 0; No information given | 1; Selected participants representative of the population they are sampled from | 1; Clear inclusion and exclusion criteria given. | 0; No justification on size of selected sample provided. | 1; Exposures of interest were measured prior to the outcome. | 0; No information given. | 1; Clear explanation of exposure of interest and how this was determined. | 0; No information given. | 1; Study clearly states diganosis of recognised hepatic pathology using eligible diagnostic modality. | 0; No information given. | 0; No information given. | 0; No information on covariates given. | 0; No adjusted/unadjusted models reported. | 8 | M |

| | | | | | | | | | | | | | | | | | | |
|---|---|---|---|---|---|---|---|---|---|---|---|---|---|---|---|---|---|---|
| Kamal, 2004 | 1; Authors clearly describe the goal of their research | 1; Clear description of the study population using demographics, location and time period. | 1; Study describes the method of selecting the participant sample. | 0; No information given | 1; Selected participants representative of the population they are sampled from | 1; Clear inclusion and exclusion criteria given. | 0; No justification on size of selected sample provided. | 1; Exposures of interest were measured prior to the outcome. | 0; No information given. | 1; Clear explanation of exposure of interest and how this was determined. | 0; No information given. | 1; Study clearly states diganosis of recognised hepatic pathology using eligible diagnostic modality. | 1; Yes, assesors were blinded to exposure status. | 0; No information given. | 0; No information on covariates given. | 0; No adjusted/unadjusted models reported. | 9 | M |
| Kamdem, 2019 | 1; Authors clearly describe the goal of their research | 1; Clear description of the study population using demographics, location and time period. | 0; Lacking details to understand how participant sampling was performed | 0; No information given | 0; Unable to determine | 1; Clear inclusion and exclusion criteria given. | 0; No justification on size of selected sample provided. | 1; Exposures of interest were measured prior to the outcome. | 0; No information given. | 1; Clear explanation of exposure of interest and how this was determined. | 0; No information given. | 1; Study clearly states diganosis of recognised hepatic pathology using eligible diagnostic modality. | 0; No information given. | 0; No information given. | 0; No information on covariates given. | 0; No adjusted/unadjusted models reported. | 6 | M |
| Leibenguth, 2024 | 1; Authors clearly describe the goal of their research | 1; Clear description of the study population using demographics, location and time period. | 1; Study describes the method of selecting the participant sample. | 0; No information given | 0; Unable to determine | 1; Clear inclusion and exclusion criteria given. | 0; No justification on size of selected sample provided. | 1; Exposures of interest were measured prior to the outcome. | 0; No information given. | 1; Clear explanation of exposure of interest and how this was determined. | 1; Assessors of exposure blinded to case or control status of participants | 1; Study clearly states diganosis of recognised hepatic pathology using eligible diagnostic modality. | 1; Assessors of outcome blinded to case or control status of participants | 0; No information given. | 0; No information on covariates given. | 0; No adjusted/unadjusted models reported. | 9 | M |

| | | | | | | | | | | | | | | | | | |
|---|---|---|---|---|---|---|---|---|---|---|---|---|---|---|---|---|---|
| Li, 1991 | 1; Authors clearly describe the goal of their research | 0; Description of the study population lacking specifics of demographics and time period | 0; Lacking details to understand how participant sampling was performed | 0; No information given | 0; Unable to determine | 0; No clear information on inclusion and exclusion criteria given. | 0; No justification on size of selected sample provided. | 1; Exposures of interest were measured prior to the outcome. | 0; No information given. | 1; Clear explanation of exposure of interest and how this was determined. | 0; No information given. | 1; Study clearly states diganosis of recognised hepatic pathology using eligible diagnostic modality. | 0; No information given. | 0; No information given. | 0; No information on covariates given. | 0; No adjusted/unadjusted models reported. | 4 | H |
| Li, 2011 | 1; Authors clearly describe the goal of their research | 0; Description of the study population lacking specifics of demographics and time period | 0; Lacking details to understand how participant sampling was performed | 0; No information given | 0; Unable to determine | 1; Clear inclusion and exclusion criteria given. | 0; No justification on size of selected sample provided. | 1; Exposures of interest were measured prior to the outcome. | 0; No information given. | 1; Clear explanation of exposure of interest and how this was determined. | 0; No information given. | 1; Study clearly states diganosis of recognised hepatic pathology using eligible diagnostic modality. | 0; No information given. | 0; No information given. | 0; No information on covariates given. | 0; No adjusted/unadjusted models reported. | 5 | H |
| Mazigo, 2017 | 1; Authors clearly describe the goal of their research | 1; Clear description of the study population using demographics, location and time period. | 1; Study describes the method of selecting the participant sample. | 0; No information given | 1; Selected participants representative of the population they are sampled from | 1; Clear inclusion and exclusion criteria given. | 1; Justification on size of selected sample provided. | 1; Exposures of interest were measured prior to the outcome. | 0; No information given. | 1; Clear explanation of exposure of interest and how this was determined. | 0; No information given. | 1; Study clearly states diganosis of recognised hepatic pathology using eligible diagnostic modality. | 0; No information given. | 0; No information given. | 0; No information on covariates given. | 1; Yes, model provided. | 10 | L |

| | | | | | | | | | | | | | | | | | | |
|---|---|---|---|---|---|---|---|---|---|---|---|---|---|---|---|---|---|---|
| Mohamed, 1998 | 1; Authors clearly describe the goal of their research | 1; Clear description of the study population using demographics, location and time period. | 0; Lacking details to understand how participant sampling was performed | 0; No information given | 0; Unable to determine | 1; Clear inclusion and exclusion criteria given. | 0; No justification on size of selected sample provided. | 1; Exposures of interest were measured prior to the outcome. | 0; No information given. | 1; Clear explanation of exposure of interest and how this was determined. | 0; No information given. | 1; Study clearly states diganosis of recognised hepatic pathology using eligible diagnostic modality. | 0; No information given. | 0; No information given. | 0; No information on covariates given. | 0; No adjusted/unadjusted models reported. | 6 | M |
| Shaala, 2015 | 1; Authors clearly describe the goal of their research | 1; Clear description of the study population using demographics, location and time period. | 0; Lacking details to understand how participant sampling was performed | 0; No information given | 0; Unable to determine | 1; Clear inclusion and exclusion criteria given. | 0; No justification on size of selected sample provided. | 1; Exposures of interest were measured prior to the outcome. | 0; No information given. | 1; Clear explanation of exposure of interest and how this was determined. | 0; No information given. | 1; Study clearly states diganosis of recognised hepatic pathology using eligible diagnostic modality. | 0; No information given. | 0; No information given. | 0; No information on covariates given. | 0; No adjusted/unadjusted models reported. | 6 | M |
| Shiha, 2002 | 1; Authors clearly describe the goal of their research | 0; Description of the study population lacking specifics of demographics and time period | 0; Lacking details to understand how participant sampling was performed | 0; No information given | 0; Unable to determine | 0; No clear information on inclusion and exclusion criteria given. | 0; No justification on size of selected sample provided. | 1; Exposures of interest were measured prior to the outcome. | 0; No information given. | 1; Clear explanation of exposure of interest and how this was determined. | 0; No information given. | 1; Study clearly states diganosis of recognised hepatic pathology using eligible diagnostic modality. | 0; No information given. | 0; No information given. | 0; No information on covariates given. | 0; No adjusted/unadjusted models reported. | 4 | H |

| | | | | | | | | | | | | | | | | | |
|---|---|---|---|---|---|---|---|---|---|---|---|---|---|---|---|---|---|
| Takagi, 2003 | 1; Authors clearly describe the goal of their research | 1; Clear description of the study population using demographics, location and time period. | 0; Lacking details to understand how participant sampling was performed | 0; No information given | 0; Unable to determine | 0; No clear information on inclusion and exclusion criteria given. | 0; No justification on size of selected sample provided. | 1; Exposures of interest were measured prior to the outcome. | 0; No information given. | 0; No clear explanation of how exposure of interest was diagnosed. | 0; No information given. | 0; Study does not provide clear explanations on the diagnostic methods used for the outcome of interest. | 0; No information given. | 0; No information given. | 0; No information on covariates given. | 0; No adjusted/unadjusted models reported. | 3 | H |
| Uchimura, 1997 | 1; Authors clearly describe the goal of their research | 1; Clear description of the study population using demographics, location and time period. | 0; Lacking details to understand how participant sampling was performed | 0; No information given | 0; Unable to determine | 0; No clear information on inclusion and exclusion criteria given. | 0; No justification on size of selected sample provided. | 1; Exposures of interest were measured prior to the outcome. | 0; No information given. | 1; Clear explanation of exposure of interest and how this was determined. | 0; No information given. | 1; Study clearly states diganosis of recognised hepatic pathology using eligible diagnostic modality. | 0; No information given. | 0; No information given. | 0; No information on covariates given. | 0; No adjusted/unadjusted models reported. | 5 | H |
| Xu, 1997 | 1; Authors clearly describe the goal of their research | 0; Description of the study population lacking specifics of demographics and time period | 0; Lacking details to understand how participant sampling was performed | 0; No information given | 0; Unable to determine | 0; No clear information on inclusion and exclusion criteria given. | 0; No justification on size of selected sample provided. | 0; Exposures of interest were measured after the outcome was established. | 0; No information given. | 1; Clear explanation of exposure of interest and how this was determined. | 0; No information given. | 0; Study does not provide clear explanations on the diagnostic methods used for the outcome of interest. | 0; No information given. | 0; No information given. | 0; No information on covariates given. | 0; No adjusted/unadjusted models reported. | 2 | H |

S – ROB score
<u>ROB score interpretation:</u>
H = high
M = moderate
L = lo

**Figure S1: Publication bias as shown by funnel plot for 33 studies included in the meta-analysis.**

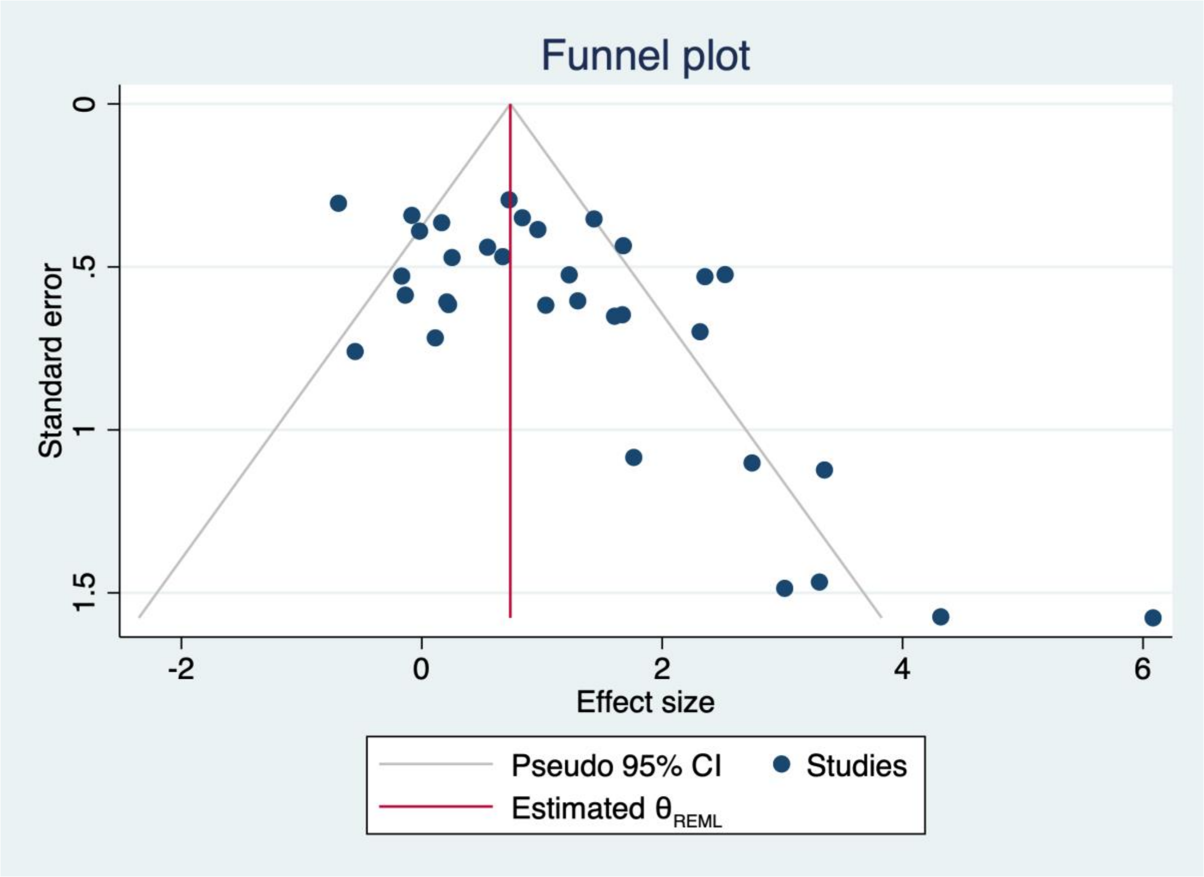

H0 beta 1 = 0
Beta1 = 2.79
SE of beta 1 = 0.606
Z = 4.60
Prob > |z| = 0.0000

### Table S13: Summary of study characteristics of all included articles

| **Characteristics** | **N** | % |
|---|---|---|
| **Hepatitis species** | N = 33 | |
| B | 5 | 15·15 |
| C | 22 | 66·67 |
| B and C | 6 | 18·18 |
| ***Schistosoma* species** | N = 33 | |
| *S.mansoni* | 24 | 72·73 |
| *S. japonicum* | 7 | 21·21 |
| *S. mekongi* | 0 | 0 |
| Combined *S. mansoni* and *S. haematobium* | 2 | 6·06 |
| **Liver disease outcomes**[*] | N = 33 | |
| Hepatomegaly | 2 | 6·06 |
| Fibrosis | 18 | 54·54 |
| Cirrhosis | 14 | 42·42 |
| Hepatocellular carcinoma | 6 | 18·18 |
| **Study aim** | N = 33 | |
| Biomarker study | 6 | 18·18 |
| Epidemiology of co-infection | 3 | 9·09 |
| Immune response to co-infection | 5 | 15·15 |
| Pathogenesis in co-infection | 14 | 42·42 |
| Treatment [†] | 2 | 6·06 |
| Unclear | 3 | 9·09 |
| **Study year** | N = 33 | |
| Before 2000 | 10 | 30·30 |
| After 2000 | 17 | 51·52 |
| Unclear | 6 | 18·18 |
| **Region** [‡] | N=33 | |
| Middle East/North Africa | 18 | 54·55 |
| Sub-Saharan Africa | 4 | 12·12 |
| East Asia/Pacific | 7 | 21·21 |
| Unclear | 4 | 12·12 |
| **Setting** [§] | N= 34 | |
| Community health centre or other village setting | 5 | 14·70 |
| Hospital | 22 | 64·70 |
| Unclear | 7 | 20·59 |
| **Sampling strategy** | N= 33 | |
| Convenience [¶] | 20 | 60·61 |
| Random sample | 3 | 9·09 |
| Unclear | 10 | 30·30 |
| **Study design** | N = 33 | |
| Cross-sectional | 21 | 63·64 |

| | | |
|---|---|---|
| Prospective cohort | 7 | 21·21 |
| Retrospective cohort | 1 | 3·03 |
| Retrospective case control | 3 | 9·09 |
| Prospective case control | 1 | 3·03 |
| **Inclusion criteria** | N = 33 | |
| Imaging findings | 1 | 3·03 |
| Positive serology | 10 | 30·30 |
| Symptoms | 1 | 3·33 |
| Other (incl. location, biopsy findings, previous treatment) | 8 | 24·24 |
| Unclear | 13 | 39·39 |
| **Exclusion criteria** | N= 33 | |
| Other liver disease present | 13 | 39·39 |
| Serology | 4 | 12·12 |
| Unclear | 16 | 48·48 |
| **Schistosoma diagnostic** [‖] | N=33 | |
| Microscopy | 22 | 66·67 |
| Serology | 18 | 54·55 |
| Imaging | 3 | 9·09 |
| Other (history, biopsy) | 13 | 39·39 |
| **Fibrosis diagnostic tools** [**] | N = 18 | |
| Biomarker | 2 | 11·11 |
| Biopsy | 8 | 44·44 |
| Imaging findings | 7 | 38·89 |
| Biopsy and Imaging | 1 | 5·56 |
| **Types of biopsy used for fibrosis diagnosis** | N = 9 | |
| Needle biopsy | 1 | 11·11 |
| Ultrasound-guided biopsy | 1 | 11·11 |
| Unclear | 7 | 77·78 |
| **Types of imaging used for fibrosis diagnosis** | N = 8 | |
| Ultrasonography | 8 | 100 |
| **Cirrhosis diagnostic tools** [††] | N=14 | |
| Biopsy | 8 | 57·14 |
| Imaging findings | 4 | 28·57 |
| Biopsy and imaging | 1 | 7·14 |
| Unclear | 1 | 7·14 |
| **Types of biopsy used for cirrhosis diagnosis** | N=9 | |
| Ultrasound-guided biopsy | 4 | 44·44 |
| Needle biopsy | 1 | 11·11 |
| Percutaneous biopsy | 1 | 11·11 |
| Unclear | 3 | 33·33 |
| **Types of imaging used for cirrhosis diagnosis** | N= 5 | |
| Ultrasonography | 5 | 100 |

| | | |
|---|---|---|
| **HCC diagnostic tool** | N=5 | |
| Biopsy | 3 | 60 |
| Imaging | 1 | 20 |
| Other (hospital records) | 1 | 20 |
| **Risk of bias** [‡‡] | N = 32 | |
| High | 14 | 443·75 |
| Moderate | 17 | 51·51 |
| Low | 1 | 3·12 |

[*]Liver disease outcomes are not mutually exclusive. For example, one study may contain two liver disease outcomes (e.g. fibrosis and cirrhosis) caused by the same hepatitis species and the same *Schistosoma* species. [†]Study aim treatment – includes studies investigating treatment efficacy in co-infection. [‡]Region definitions using World Bank country regions (https://www.worldbank.org/en/where-we-work). [§]One study (Xu 1997)[41] was set in both urban and rural settings, and so is included in both categories. [¶]Convenience sample = study participants sampled from patient group attending health care facility in any capacity. [||]Schistosoma diagnostic methods are not mutually exclusive i.e. multiple diagnostic methods used within the same study. ** Fibrosis diagnostic tools are not mutually exclusive i.e. multiple methods used within the same study. [††]Cirrhosis diagnostic tools are not mutually exclusive i.e. multiple methods used within the same study. [‡‡]One study (Houlihan et al., 2011) excluded from risk of bias assessment, as only conference abstract available.

**Figure S2: Pooled effect sizes for any liver pathology with outliers removed**

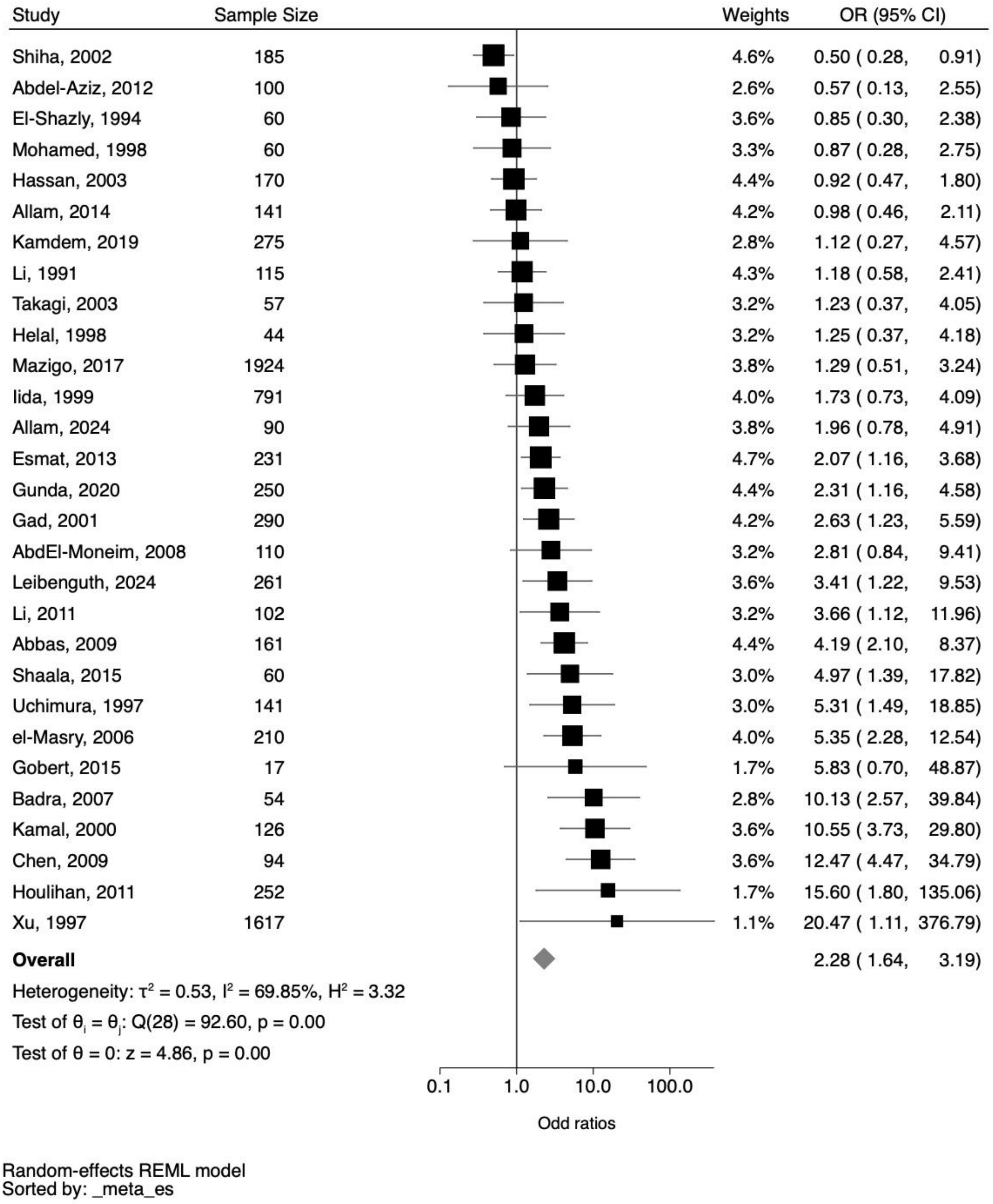

Outliers were defined as studies with an OR >25. The studies with outliers were Kamal et al. 2001, Kamal et al. 2004 and Haleem et al. 2015.

**Figure S3: Influence of any schistosome and hepatitis B or C co-infection on the presence of liver fibrosis.**

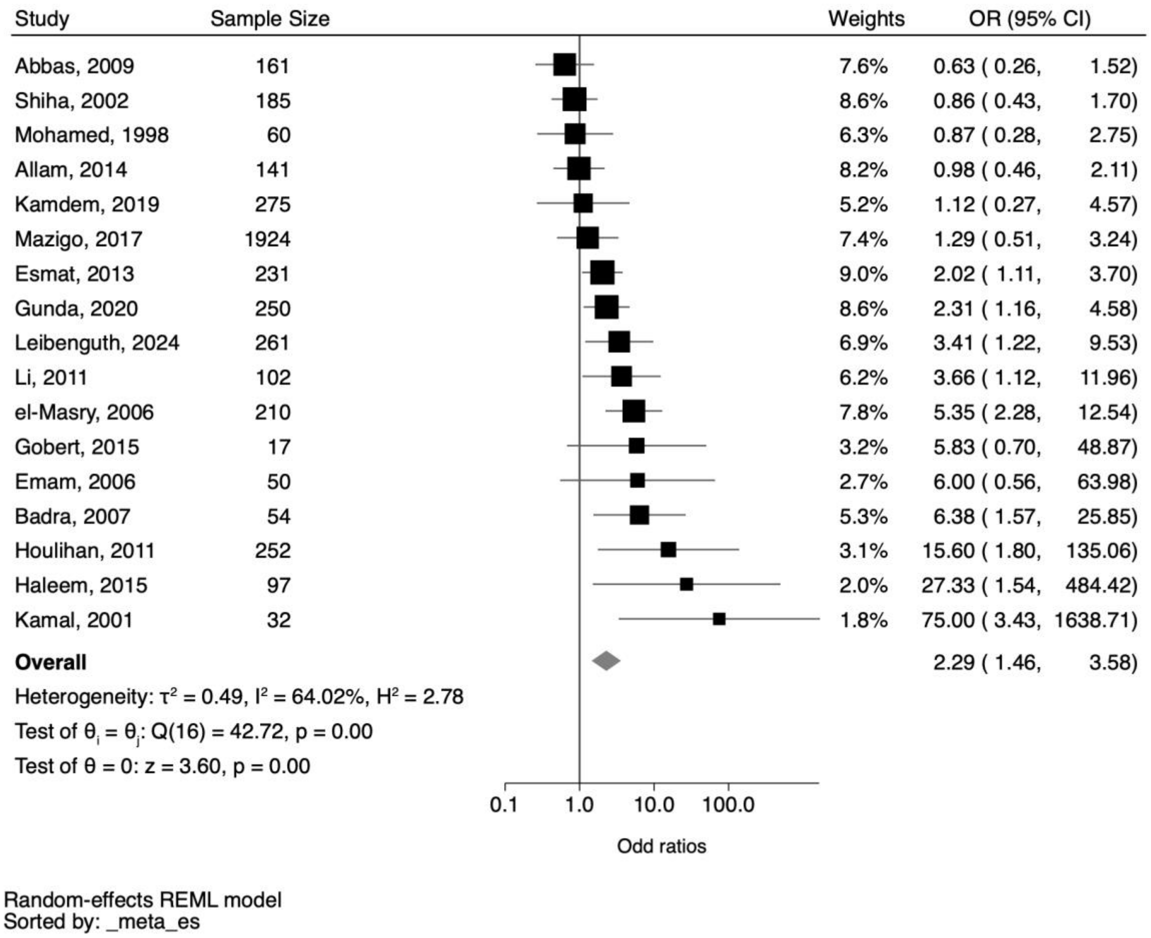

The pooled effect sizes are based on 18 studies reporting on fibrosis outcomes. OR (Odds Ratio), 95% CI (95% Confidence Interval), $I^2$ = statistic of heterogeneity.

**Figure S4: Pooled effect sizes for fibrosis with outliers removed**

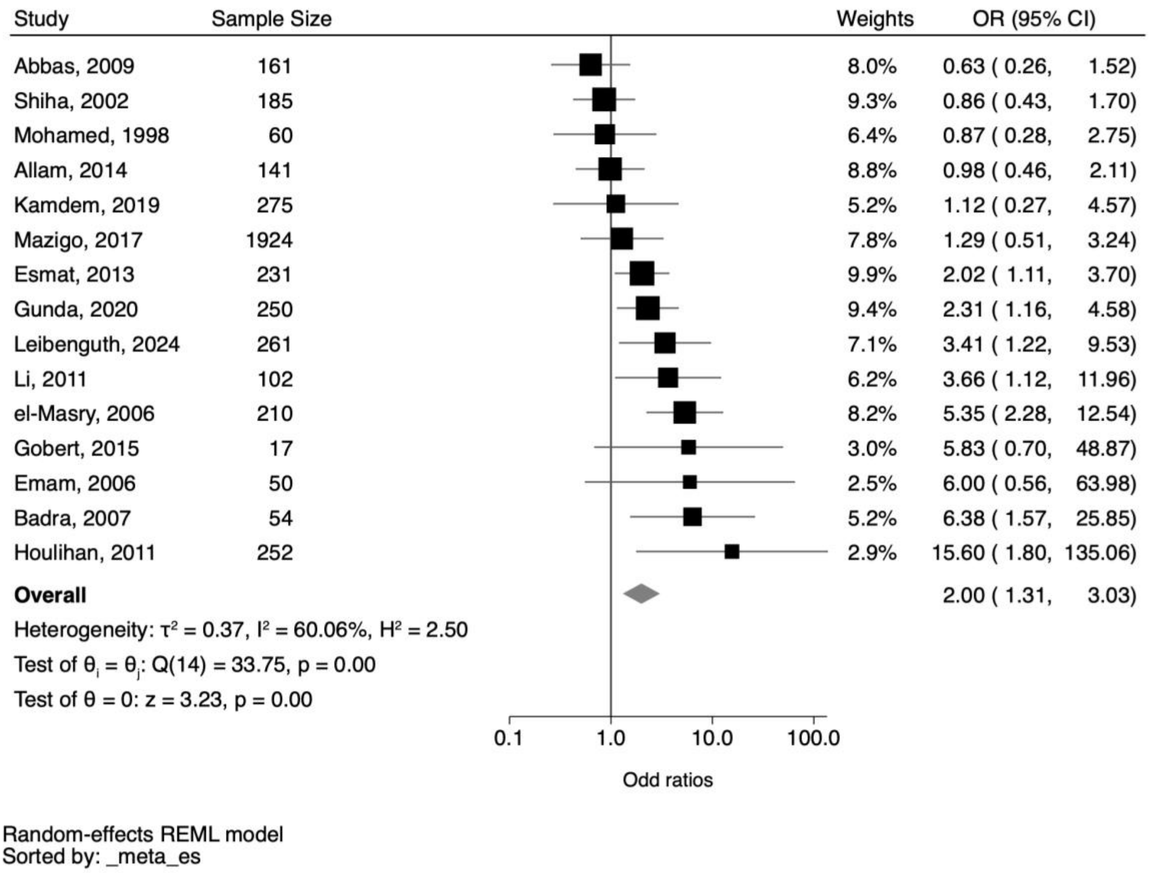

Outliers were defined as studies with an OR >25. The studies with outliers were Kamal et al. 2001, Kamal et al. 2004 and Haleem et al. 2015.

**Figure S5: Influence of any schistosome and hepatitis B or C co-infection on the presence of liver cirrhosis.**

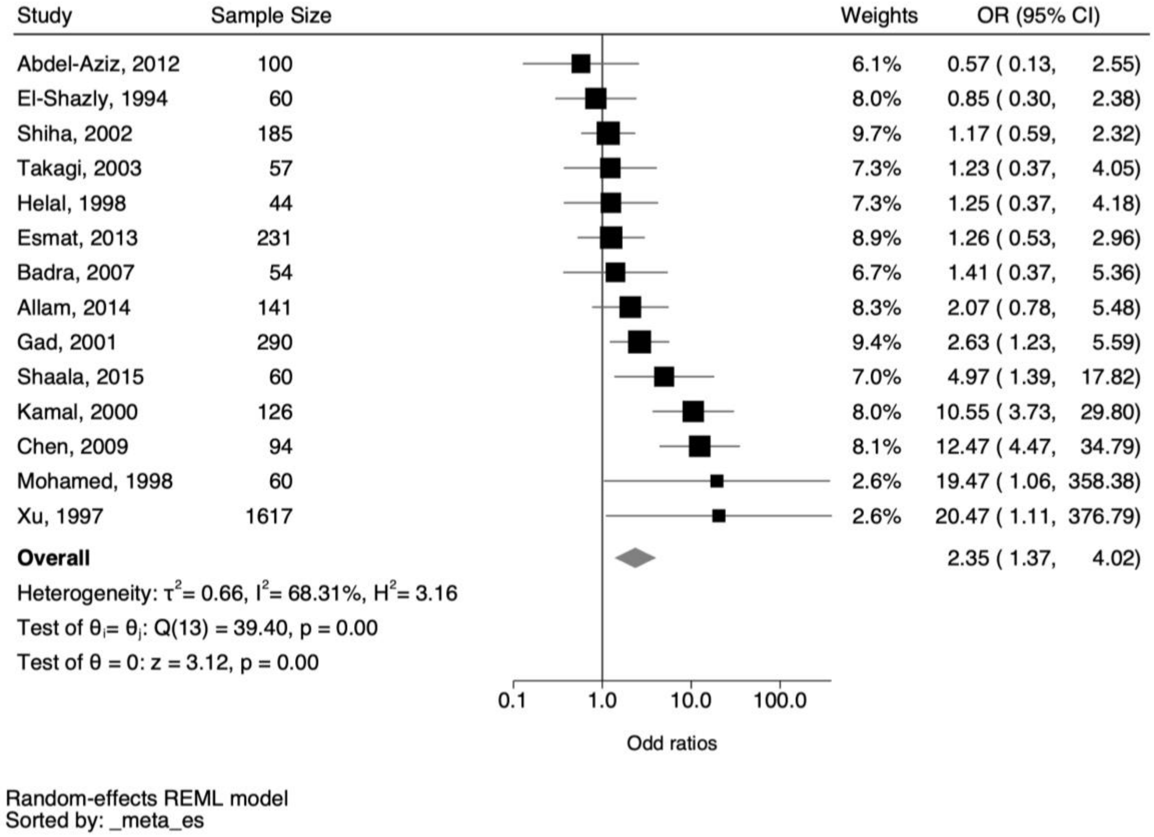

The pooled effect sizes are based on 14 studies reporting on cirrhosis outcomes. OR (Odds Ratio), 95% CI (95% Confidence Interval), $I^2$ = statistic of heterogeneity.

**Figure S6: Influence of any schistosome and hepatitis B or C co-infection on the presence of hepatocellular carcinoma**

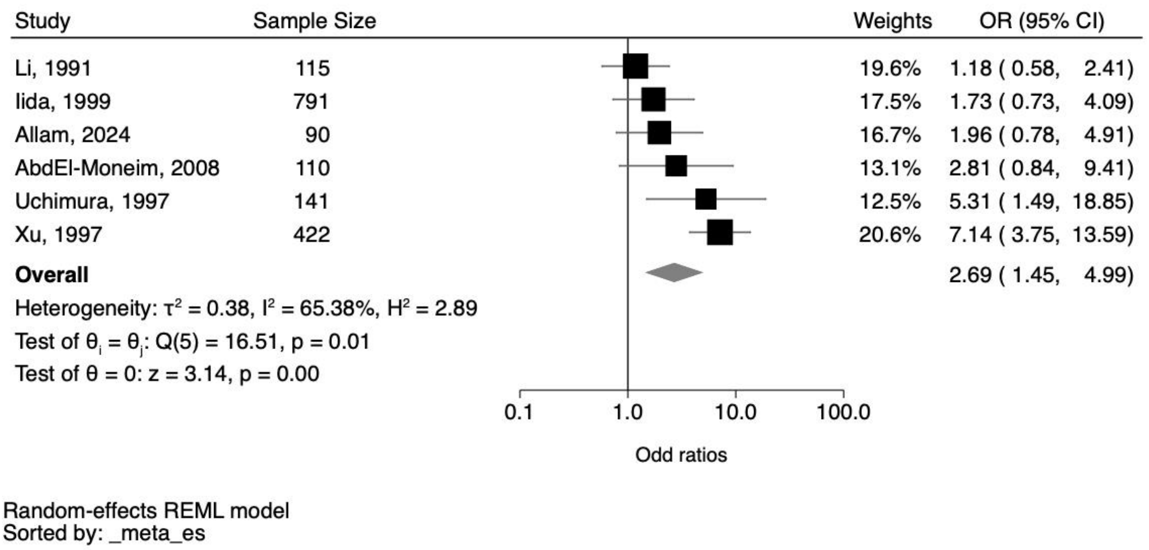

The pooled effect sizes are based on 6 studies reporting on hepatocellular carcinoma outcomes. OR (Odds Ratio), 95% CI (95% Confidence Interval), $I^2$ = statistic of heterogeneity.

**Table S14: Pooled effect sizes for all liver pathology outcomes with 10 studies removed that included healthy controls as reference population**

| Pathology | Number of studies | Number of participants with pathology | Number of co-infected individuals | OR (95% CI) | $I^2$ |
|---|---|---|---|---|---|
| Any pathology | 23 | 5054 | 910 | 2·63 (1·62 – 4·25) | 79·16% |
| Liver fibrosis | 12 | 3376 | 482 | 2·22 (1·29 – 3·81) | 65·02% |
| Cirrhosis | 12 | 1212 | 491 | 2·19 (1·20 – 3·97) | 70·62% |
| Hepatocellular carcinoma | 4 | 1137 | 183 | 1·81 (1·11 – 2·96) | 16·59% |

Studies removed were Abbas, 2009; Abd El-Moneim, 2008; el-Masry, 2006; Gad, 2001; Gobert, 2015; Kamdem, 2019; Leibenguth, 2024; Xu, 1997; Hassan, 2003 and Emam, 2006

### Text S2: Subgroup variables

Regions were classified using World Bank definitions of regions. These were categorically coded as East Asia/Pacific, South Asia, Europe/Central Asia, Latin America/Caribbean, Middle East/North Africa, Africa. There were only enough studies in the East Asia/Pacific and Middle East/North Africa region to perform subgroup analyses.

Hepatitis species were extracted based on author-reported diagnostic methods that were eligible for inclusion. Hepatitis species was coded as B, B and C or C.

*Schistosoma* species were extracted based on author-reported diagnostic methods that were eligible for inclusion: *S. mansoni, S. japonicum, S. mekongi* and Combined S. mansoni and S. haematobium. There were only enough studies in the *S. mansoni, S. japonicum* group to perform subgroup analyses.

Study aims were grouped using either author-definitions of their aim or assigned based on the outcome of the study. The aims were categorically grouped into one of five overarching aims: biomarker study, epidemiology of co-infection, immune response to co-infection, pathogenesis in co-infection and treatment.

Diagnostic tools for fibrosis and cirrhosis were classified using author-reported diagnostic modalities that were eligible for inclusion: biopsy, imaging, biomarker, combined biopsy and imaging.

**Table S15: Subgroup analyses for liver fibrosis caused by co-infection.**

| Subgroups* | Studies | Number of participants | Number of co-infected individuals | Number of individuals with fibrosis | OR (95% CI) | $I^2$ | Test of group differences |
|---|---|---|---|---|---|---|---|
| **World Bank Region** † | | | | | | | |
| Middle East/ North Africa | 8 | 979 | 375 | 445 | 1·51 (0·83 – 2·73) | 62·02% | Qb(1) = 0·55, p= 0·46 |
| Sub-Saharan Africa | 4 | 2710 | 140 | 318 | 2·00 (1·26 – 3·16) | 0·00% | |
| **Hepatitis species** | | | | | | | |
| B | 3 | 519 | 54 | 133 | 4·15 (1·31 – 13·09) | 40·50% | Qb(2) = 1·17, p= 0·56 |
| C | 11 | 3145 | 507 | 634 | 2·05 (1·10 – 3·83) | 73·82% | |
| B and C | 3 | 638 | 98 | 289 | 2·69 (1·36 – 5·31) | 0·00% | |
| **Diagnostic tool** ‡ | | | | | | | |
| Biopsy | 7 | 810 | 294 | 406 | 3·12 (1·02 –9·54) | 83·58% | Qb(1) = 0·92, p=0·34 |
| Imaging | 7 | 2930 | 212 | 329 | 1·69 (0·95 – 2·99) | 37·14% | |
| **Risk of bias** § | | | | | | | |
| Medium/Low | 11 | 3272 | 395 | 590 | 1·99 (1·13 – 3·49) | 59·72% | Qb(1) = 0·19, p= 0·66 |
| High | 5 | 778 | 257 | 430 | 2·45 (1·16 – 5·20) | 69·28% | |
| **Study aim** ‖ | | | | | | | |
| Biomarker | 4 | 632 | 179 | 387 | 4·13 (1·39 – 12·34) | 44·48% | Qb(2) = 1·28, p= 0·53 |
| Immune response | 3 | 334 | 140 | 121 | 2·44 (0·19 – 30·64) | 93·00% | |
| Pathogenesis | 6 | 893 | 239 | 392 | 2·01 (1·09 – 3·69) | 60·73% | |

*One study (Kamal et al. 2004) removed from all subgroups due to a zero value. †Two studies (Gobert, 2015 and Li, 2011) set in the East Asia/Pacific region removed from this subgroup analysis. ‡ One study using biomarkers as a diagnostic tool for fibrosis (el-Masry, 2006) and one study using both biopsy and imaging (Li, 2011) removed from this subgroup analysis. § study (Houlihan, 2011) removed from this subgroup. || One study with the aim "epidemiology" (Mazigo, 2017) and three studies with unclear aim (Gobert, 2015; Gunda, 2020; Houlihan, 2011) were removed from this subgroup analysis.

**Table S16: Subgroup analyses for cirrhosis caused by co-infection.**

| Subgroups | Studies | Number of participants | Number of co-infected individuals | Number of individuals with cirrhosis | OR (95% CI) | $I^2$ | Test of group differences |
|---|---|---|---|---|---|---|---|
| **Diagnostic tool** | | | | | | | |
| Biopsy | 8 | 828 | 314 | 208 | 1·74 (0·87 – 3·49) | 70·81% | Qb (1) = 0·34, p= 0·56 |
| Imaging | 4 | 548 | 174 | 95 | 2·26 (1·34 – 3·82) | 0·00% | |

**Figure S7: Pooled effect sizes for any liver pathology with high RoB studies removed**

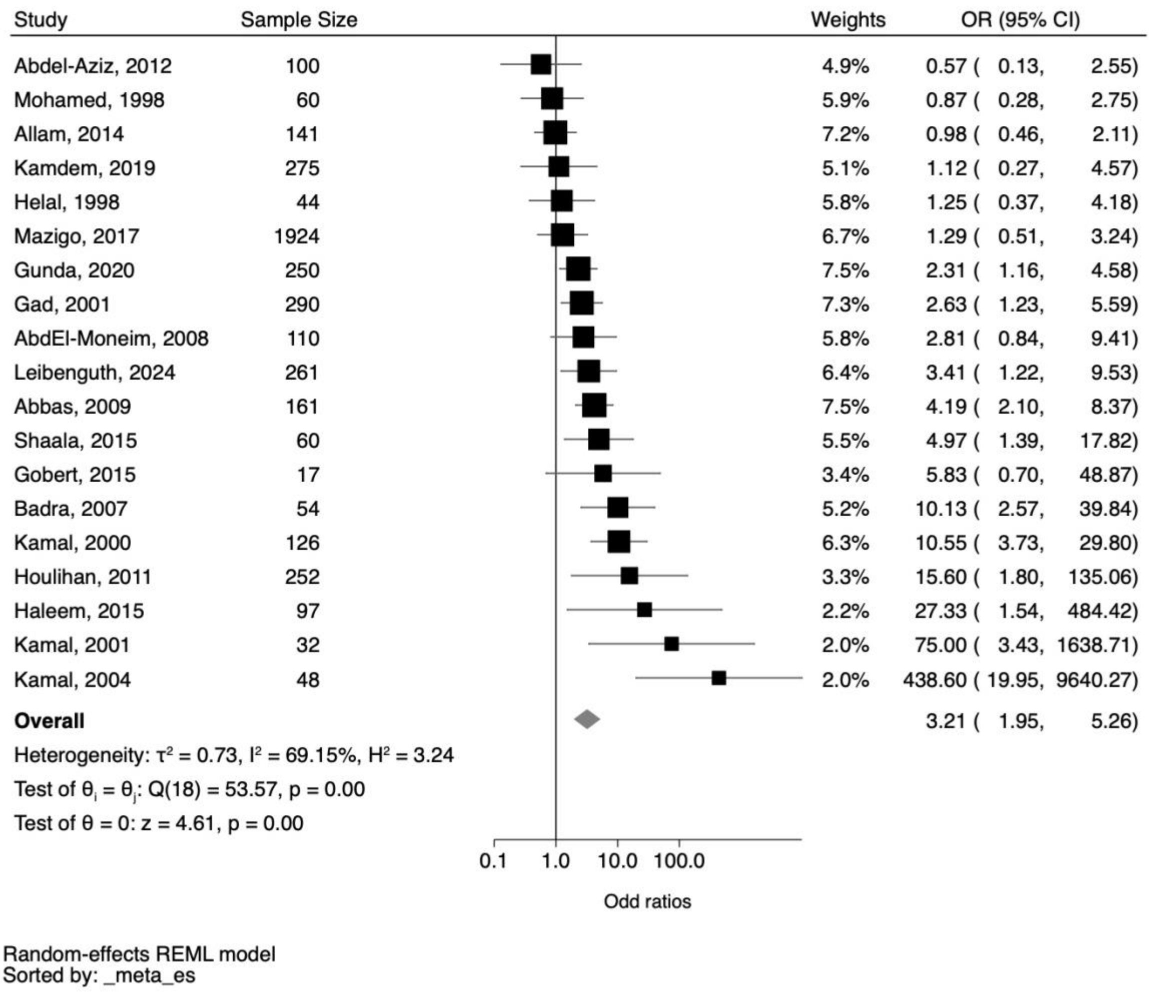

Studies removed were Shiha, 2002; El-Shazly, 1994; Hassan, 2003; Li, 1991; Takagi, 2003; Iida, 1999; Allam, 2024; Esmat, 2013; Li, 2011; Uchimura, 1997; el-Masry, 2006; Chen, 2009; Xu, 1997 and Emam, 2006.

### Figure S8: Pooled effect sizes for fibrosis with high RoB studies removed

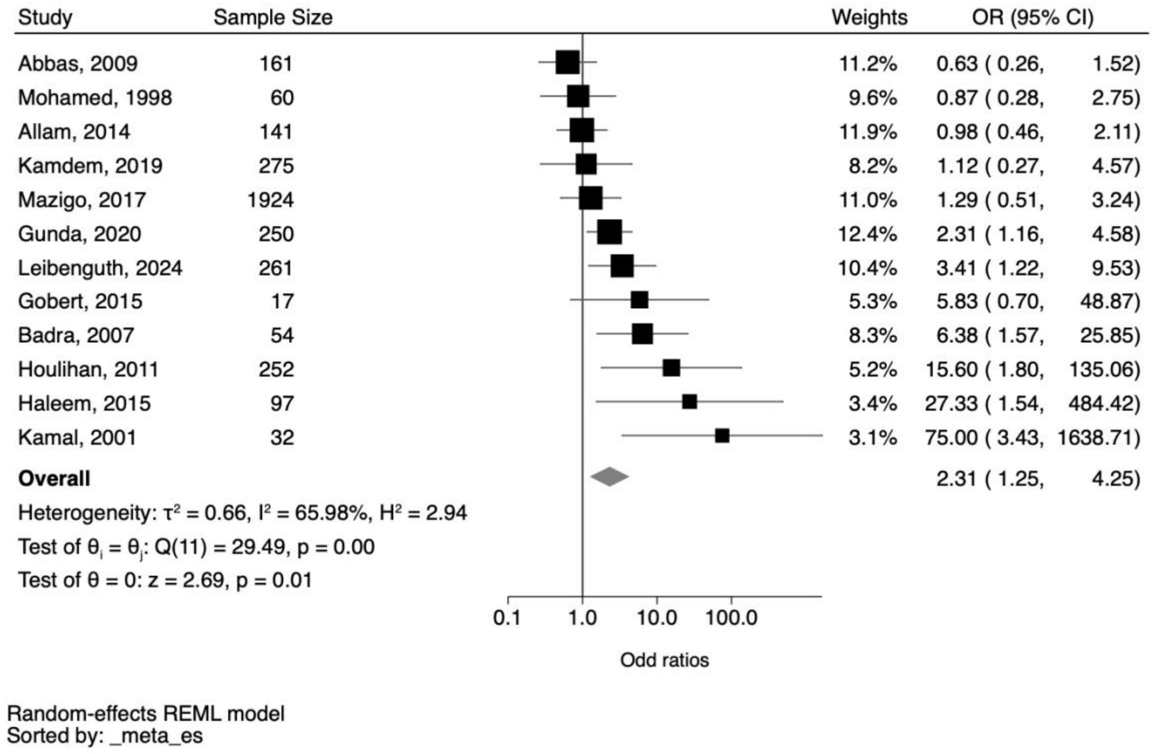

Studies removed were Shiha, 2002; Esmat, 2013; Li, 2011; el-Masry, 2006; Emam, 2006.

#### Figure S9: Pooled effect sizes for cirrhosis with high RoB studies removed

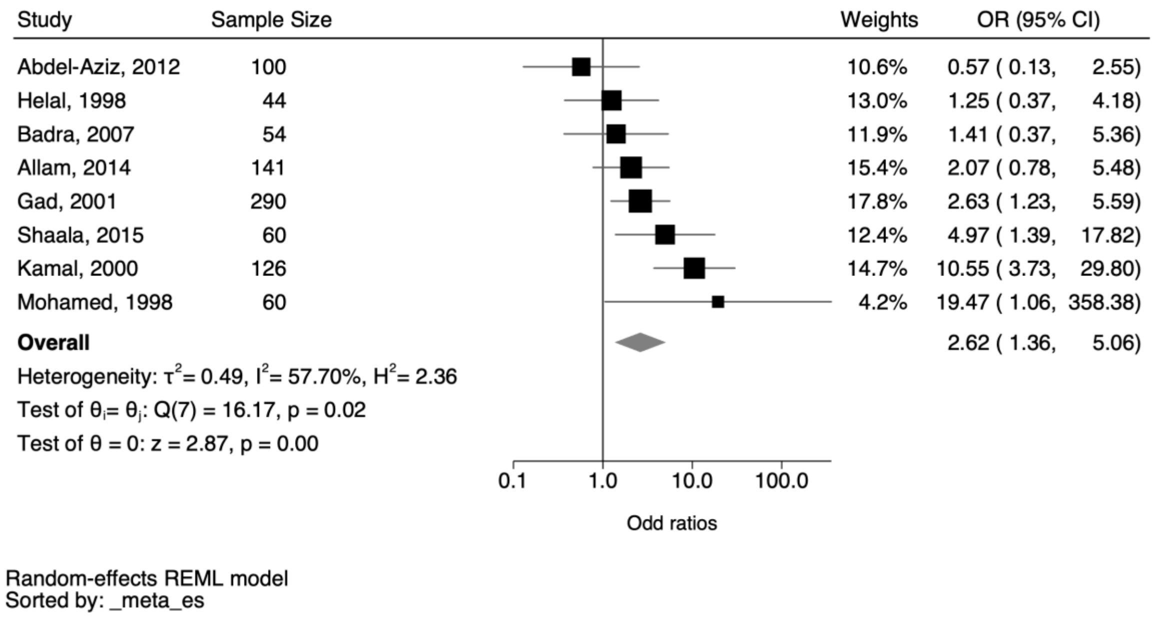

Studies removed were El-Shazly, 1994; Shiha, 2002; Takagi, 2003; Esmat, 2013; Chen, 2009 and Xu, 1997.
